# A co-infection model for Oncogenic HPV and TB with Optimal Control and Cost-Effectiveness Analysis

**DOI:** 10.1101/2020.09.15.20195297

**Authors:** A. Omame, D. Okuonghae

## Abstract

A co-infection model for oncogenic Human papillomavirus (HPV) and Tuberculosis (TB), with optimal control and cost-effectiveness analysis is studied and analyzed to assess the impact of controls against incident infection and against infection with HPV by TB infected individuals as well as optimal TB treatment in reducing the burden of the co-infection of the two diseases in a population. The co-infection model is shown to exhibit the dynamical property of backward bifurcation when the associated reproduction number is less than unity. Furthermore, it is shown that TB and HPV re-infection parameters (*ϕ*_p_ = 0 and *σ*_t_ = 0) as well as TB exogenous re-infection term (*ε*_1_ 0) induced the phenomenon of backward bifurcation in the oncogenic HPV-TB co-infection model. The global asymptotic stability of the disease-free equilibrium of the co-infection model is also proven **not to exist**, when the associated reproduction number is below unity. The necessary conditions for the existence of optimal control and the optimality system for the co-infection model is established using the Pontryagin ‘s Maximum Principle. Uncertainty and global sensitivity analysis are also carried out to determine the top ranked parameters that drive the dynamics of the co-infection model, when the associated reproduction numbers as well as the infected populations are used as response functions. Numerical simulations of the optimal control model reveal that the intervention strategy which combines and implements control against HPV infection by TB infected individuals as well as TB treatment control for dually infected individuals is the most cost-effective of all the control strategies for the control and management of the burden of oncogenic HPV and TB co-infection.

## 1 Introduction

Tuberculosis (TB), caused by *Mycobacterium tuberculosis* (MTB), remains a global dominant cause of mortality and morbidity, although treatment and preventive measures are available [24, 38]. Globally, TB is one of the dominant cause of mortality from a single infectious disease agent, responsible for roughly 40% of deaths [36]. Worldwide, approximately, 10 million new cases of TB and about 1.57 million TB-induced deaths were reported in 2017, with incidence and mortality highest in South-Eastern Asia and Africa[36]. Estimates have equally shown that about one-fourth of the world ‘s population is infected with latent TB and is at the risk of progression to active TB [15].

Cancer is the major cause of mortality in developed countries in Europe and America and the second leading cause of death in developing countries [12]. Epidemiological evidences have proven that 80% of global cancer cases are attributed to the oncogenic Human papillomavirus (HPV) [2]. In a study carried out by Zetola *et al*. [37], it was revealed that prior TB infection is mostly common among patients with cases of cervical cancer. Likewise, Zhao *et al* [39], in another study, showed that TB infection is associated with high vulnerability to oncogenic HPV infection. According to Zetola *et al*. [37], oncogenic HPV and tuberculosis always co-exist, and the immune suppression caused by cancer (which is a consequence of oncogenic HPV) can result in latent TB re-activation, hence, leading to high mortality. In addition, persistent gynecologic TB infection, resulting in chronic inflammation, could be a high risk factor in the progression of oncogenic HPV infection to cervical cancer [37, 39].

Mathematical models have recently been studied to understand the transmission dynamics of human papillomavirus infection (See Omame et al. [23] and the references included therein). Malik *et al*. [17] investigated an optimal control model for HPV, incorporating optimal vaccination strategies for females in the population. In particular, they considered when three vaccines are used at the same time in comparison to the case where the bivalent *cervarix* and quadrivalent *Gardasil 4* vaccines were used at the initial stage and then, during the course of the vaccination program, one or two of them are interchanged with the nonavalent *Gardasil 9* vaccine. Saldana *et al*. [29] developed and analyzed a two-sex model of HPV with optimal control analysis, incorporating vaccination of adolescents, adults, and screening. They showed that to optimally curb the spread of HPV in a population, vaccination should be administered both before and after the commencement of active sexual life for both females and males in the population. Likewise, TB only models have been extensively studied in the literature (see for instance, [10, 21, 22, 33] and many others).

Lately, mathematical models have been developed to consider the optimal control strategies for the dynamics of infectious diseases including their co-infections [1, 9, 11, 20, 26, 27, 31, 32, 34]. Agusto and Adekunle [1] studied the optimal control and cost-effectiveness analysis of the co-infection of drug-resistant tuberculosis and HIV/AIDS. They considered different control strategies for the control of the co-infections of both diseases. They showed that the strategy which combines efforts on minimizing the number of individuals with drug-sensitive and drug-resistant TB and case finding, prevention of treatment failure in the drug-sensitive TB infectious individuals and the treatment of individuals with drug-resistant TB is the most cost-effective in curbing the co-infections of drug-resistant TB and HIV/AIDS. Okosun and Makinde [20] considered a malaria-cholera co-infection model, in the presence of prevention and treatment controls for both diseases. Applying the Pontryagin ‘s Maximum principle, they derived the necessary conditions for optimality and equally showed using simulations, that to successfully curb malaria spread, control strategies must include cholera intervention strategies as well. Tilahun *et al*. [32] investigated a Pneumonia-Typhoid co-infection model with cost-effective optimal control analysis, incorporating preventive and treatment controls for both diseases. They showed that the strategy that combines Pneumonia treatment and Typhoid fever prevention is the most cost-effective in reducing the burden of the co-infections of Pnemonia and Typhoid fever. The researchers in [9] analyzed an optimal control model for HIV and TB co-infection, capturing resistance of HIV to anti-retroviral therapy. The authors showed that a combination of TB treatment and anti-retroviral therapy for HIV is the most effective in reducing the burden of the co-infection of the two diseases. In addition, Tanvi and Aggarwal [31] studied an HIV-TB co-infection, incorporating detection and treatment for both diseases. They observed that the strategy that implements detection for both diseases yielded the most economic and epidemic gains infighting the co-infections of the two diseases.

To the best of the authors ‘knowledge, optimal control analysis has never been applied to the co-infection of Human papillomavirus and TB in the literature. Hence, it will be imperative to investigate the optimal control and cost-effectiveness analysis of the co-infections of oncogenic HPV and TB. In particular, this paper extends the work of Omame *et al*. [24] *by* assessing the impact of HPV prevention and TB treatment controls on the control and management of the co-infections of the two diseases.

The rest of the paper is organized as follows. The model is formulated in Section 2, alongside the basic properties of the model. The co-infection model without controls is analyzed qualitatively in Section 3. The optimal control model is considered in Section 4. Simulations of the model are carried out in Section 5 while Section 6 gives the concluding remarks.

## 2 Model formulation and basic properties of the model

The total sexually active population at time *t*, denoted by *N*_H_(*t*), is divided into eleven mutually-exclusive compartments: Susceptible individuals (*S*(*t*)), individuals infected with HPV (*I*_HP_ (*t*)), individuals who have recovered from or cleared HPV infection (*R*_HP_ (*t*)), individuals with persistent HPV infection (*P*_HP_ (*t*)), individuals with latent TB (*E*_T_(*t*)), individuals with active TB infection (*A*_T_(*t*)), individuals treated of TB (*T*_T_(*t*)), individuals dually infected with HPV and latent 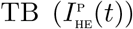, individuals dually infected with HPV and active 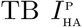, individuals dually infected with persistent HPV and latent TB, 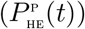 individuals dually infected with persistent HPV and active 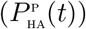. Thus

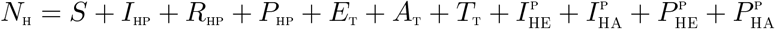

Based on the above formulations and assumptions, the HPV-TB Coinfection model is given by the following deterministic system of non-linear differential equations (flow diagram of the model is shown in Figure 1, the associated state variables and parameters are well described in Tables 1 and 2):

**Table 1:**
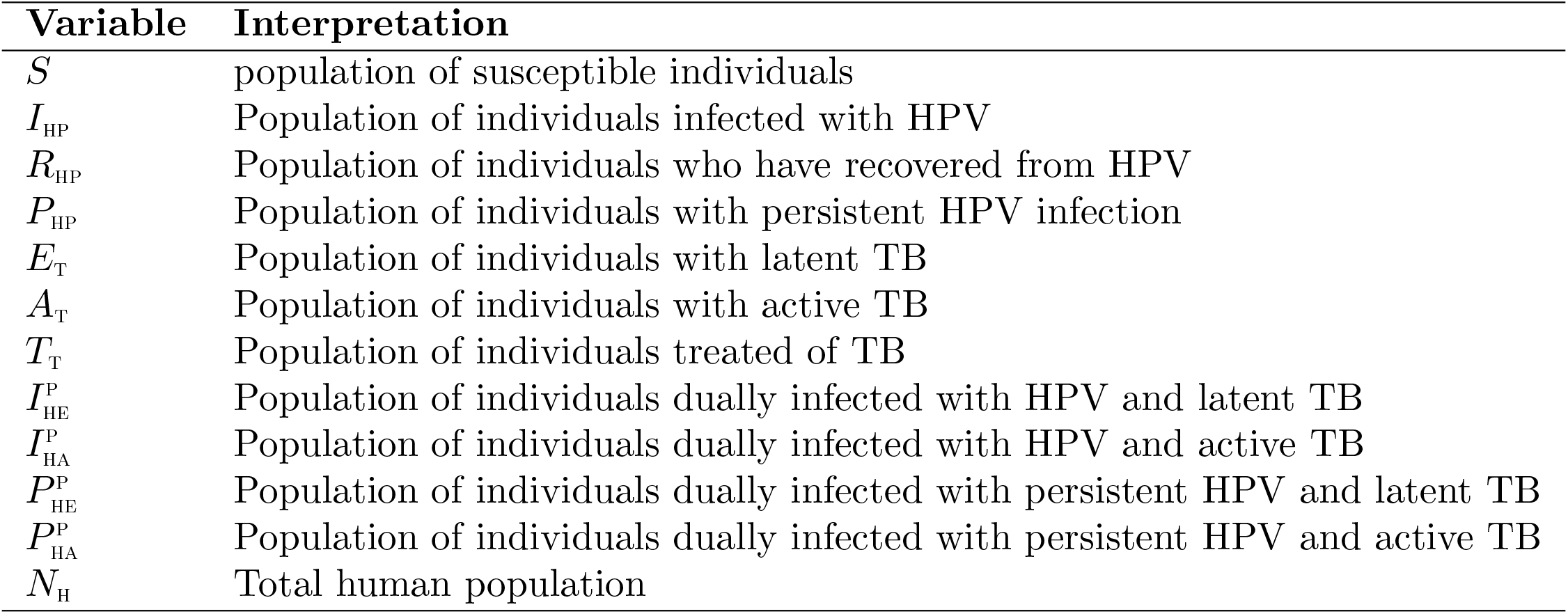
Description of variables in the model (1).

| Variable | Interpretation |
| --- | --- |
| $S$ | population of susceptible individuals |
| $I_{\text{HP}}$ | Population of individuals infected with HPV |
| $R_{\text{HP}}$ | Population of individuals who have recovered from HPV |
| $P_{\text{HP}}$ | Population of individuals with persistent HPV infection |
| $E_{\text{T}}$ | Population of individuals with latent TB |
| $A_{\text{T}}$ | Population of individuals with active TB |
| $T_{\text{T}}$ | Population of individuals treated of TB |
| $I_{\text{HE}}^{\text{p}}$ | Population of individuals dually infected with HPV and latent TB |
| $I_{\text{HA}}^{\text{p}}$ | Population of individuals dually infected with HPV and active TB |
| $P_{\text{HE}}^{\text{p}}$ | Population of individuals dually infected with persistent HPV and latent TB |
| $P_{\text{HA}}^{\text{p}}$ | Population of individuals dually infected with persistent HPV and active TB |
| $N_{\text{H}}$ | Total human population |

**Table 2:**
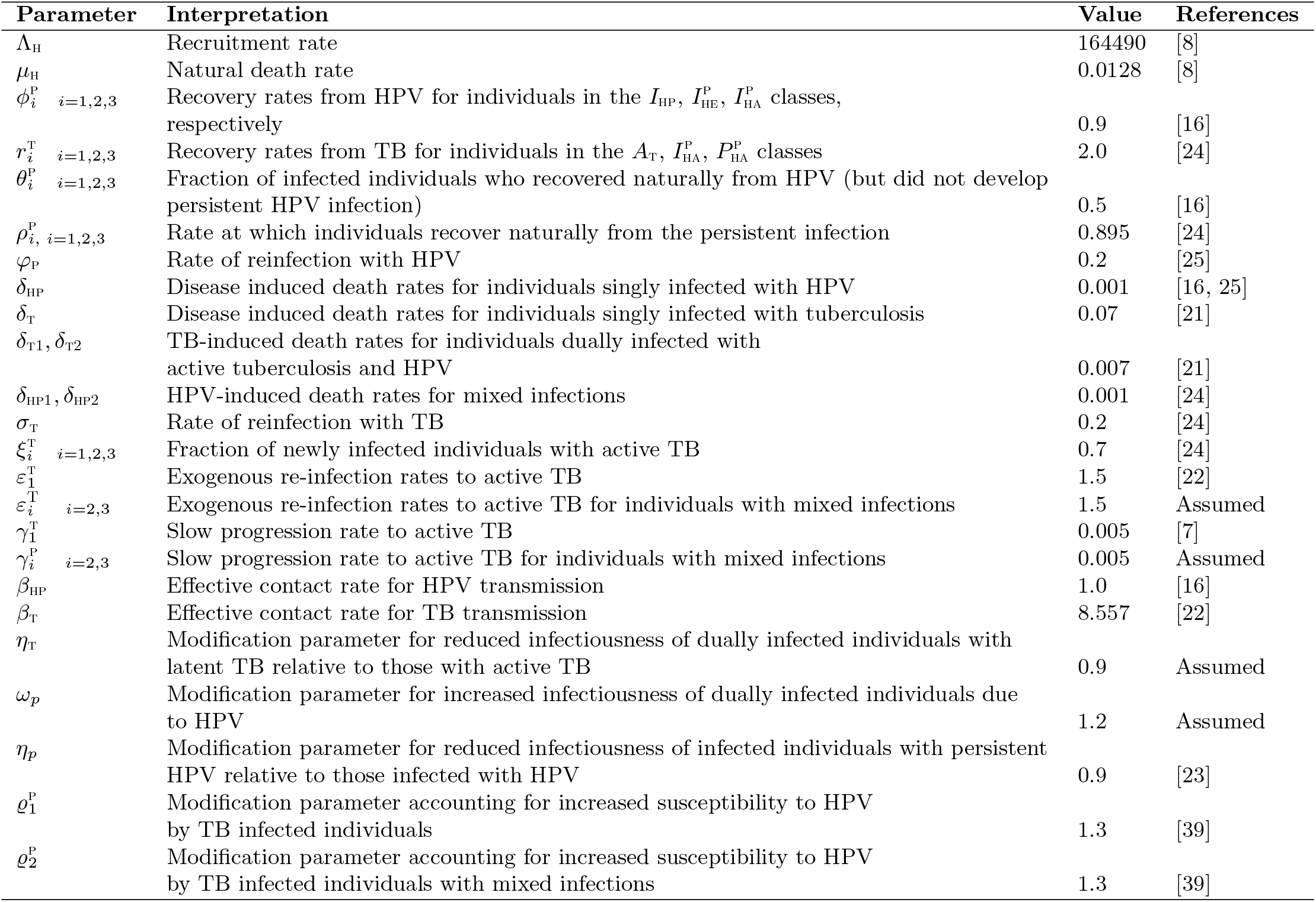
Description of parameters in the model (1).

| Parameter | Interpretation | Value | References |
| --- | --- | --- | --- |
| $\Lambda_H$ | Recruitment rate | 164490 | [8] |
| $\mu_H$ | Natural death rate | 0.0128 | [8] |
| $\phi_i^P$ $i=1,2,3$ | Recovery rates from HPV for individuals in the $I_{HP}$, $I_{HE}^P$, $I_{HA}^P$ classes, respectively | 0.9 | [16] |
| $r_i^T$ $i=1,2,3$ | Recovery rates from TB for individuals in the $A_T$, $I_{HA}^P$, $P_{HA}^P$ classes | 2.0 | [24] |
| $\theta_i^P$ $i=1,2,3$ | Fraction of infected individuals who recovered naturally from HPV (but did not develop persistent HPV infection) | 0.5 | [16] |
| $\rho_i^P$ $i=1,2,3$ | Rate at which individuals recover naturally from the persistent infection | 0.895 | [24] |
| $\varphi_P$ | Rate of reinfection with HPV | 0.2 | [25] |
| $\delta_{HP}$ | Disease induced death rates for individuals singly infected with HPV | 0.001 | [16, 25] |
| $\delta_T$ | Disease induced death rates for individuals singly infected with tuberculosis | 0.07 | [21] |
| $\delta_{T1}, \delta_{T2}$ | TB-induced death rates for individuals dually infected with active tuberculosis and HPV | 0.007 | [21] |
| $\delta_{HP1}, \delta_{HP2}$ | HPV-induced death rates for mixed infections | 0.001 | [24] |
| $\sigma_T$ | Rate of reinfection with TB | 0.2 | [24] |
| $\xi_i^T$ $i=1,2,3$ | Fraction of newly infected individuals with active TB | 0.7 | [24] |
| $\varepsilon_1^T$ | Exogenous re-infection rates to active TB | 1.5 | [22] |
| $\varepsilon_i^T$ $i=2,3$ | Exogenous re-infection rates to active TB for individuals with mixed infections | 1.5 | Assumed |
| $\gamma_1^T$ | Slow progression rate to active TB | 0.005 | [7] |
| $\gamma_i^P$ $i=2,3$ | Slow progression rate to active TB for individuals with mixed infections | 0.005 | Assumed |
| $\beta_{HP}$ | Effective contact rate for HPV transmission | 1.0 | [16] |
| $\beta_T$ | Effective contact rate for TB transmission | 8.557 | [22] |
| $\eta_T$ | Modification parameter for reduced infectiousness of dually infected individuals with latent TB relative to those with active TB | 0.9 | Assumed |
| $\omega_P$ | Modification parameter for increased infectiousness of dually infected individuals due to HPV | 1.2 | Assumed |
| $\eta_P$ | Modification parameter for reduced infectiousness of infected individuals with persistent HPV relative to those infected with HPV | 0.9 | [23] |
| $\varrho_1^P$ | Modification parameter accounting for increased susceptibility to HPV by TB infected individuals | 1.3 | [39] |
| $\varrho_2^P$ | Modification parameter accounting for increased susceptibility to HPV by TB infected individuals with mixed infections | 1.3 | [39] |

**Figure 1:**
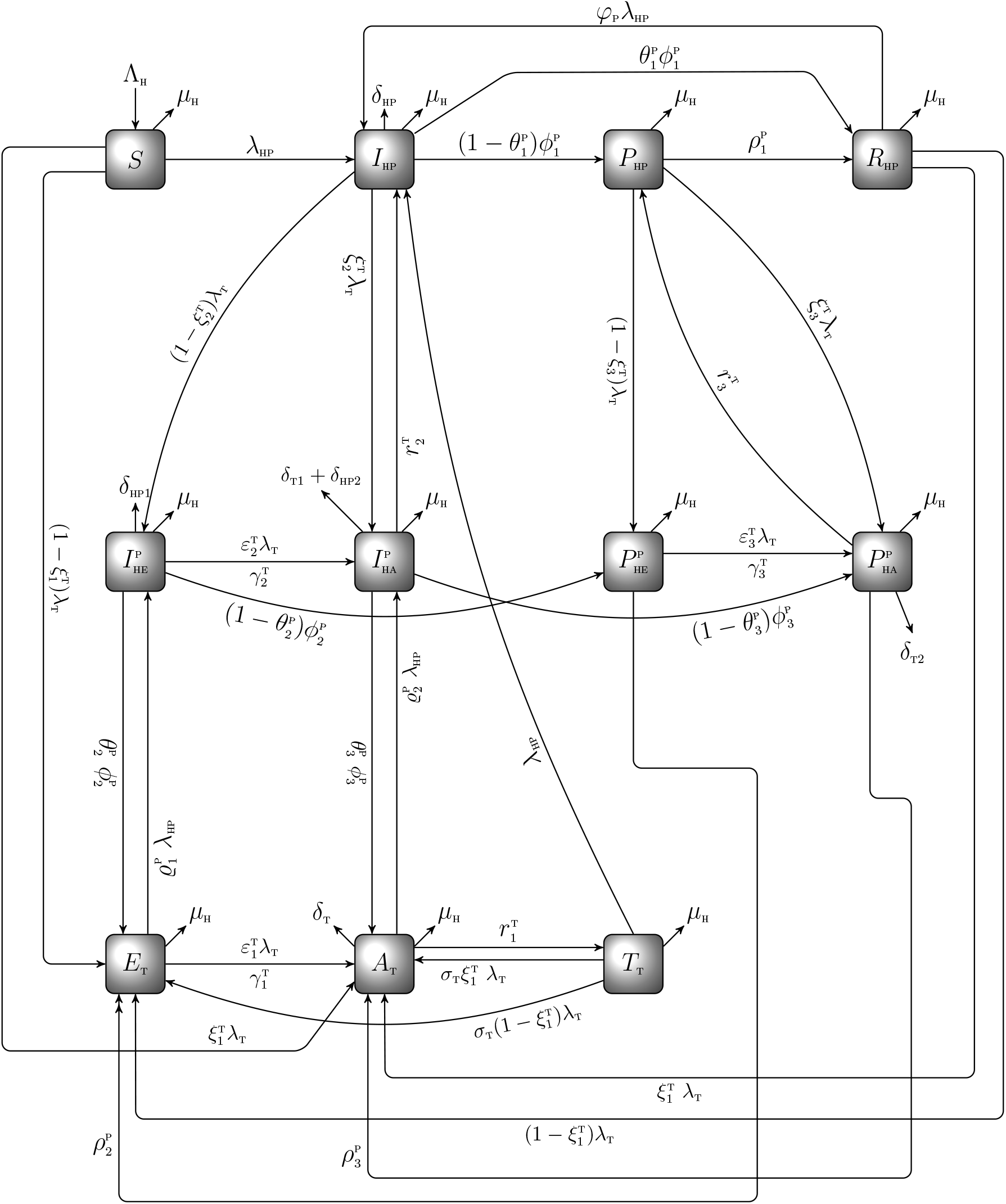
Schematic diagram of the model (1)

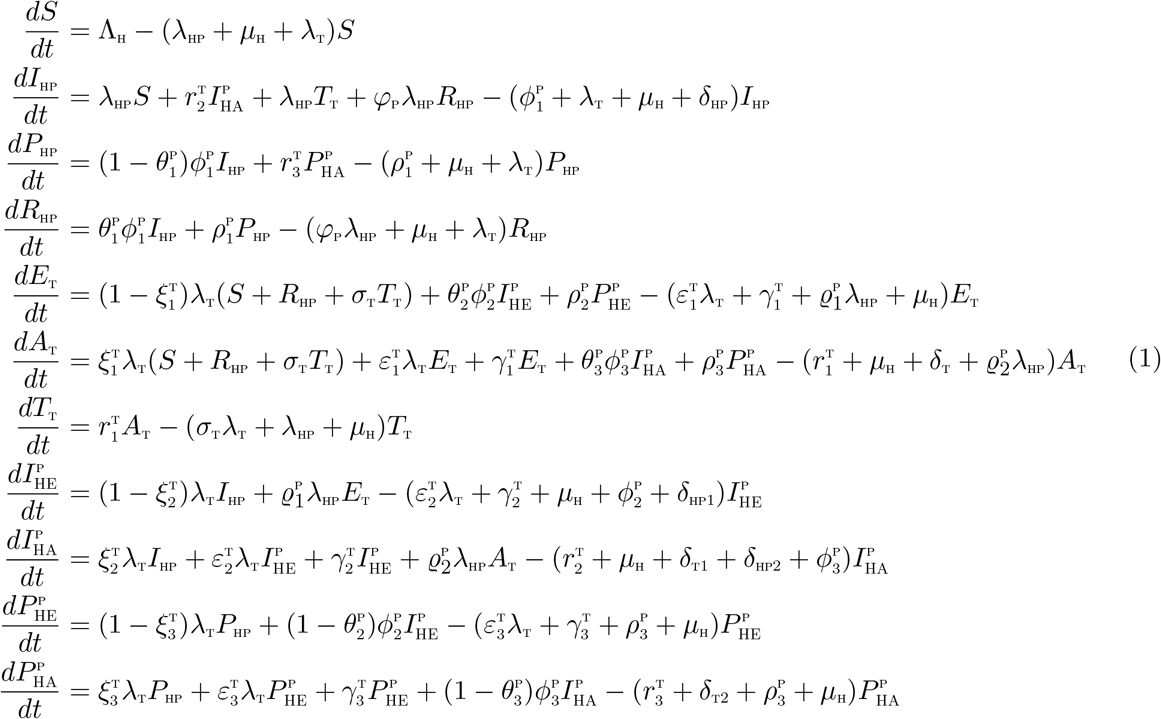

where:

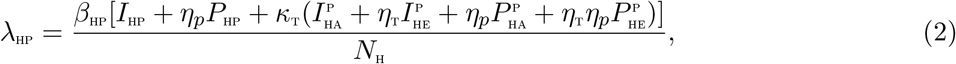

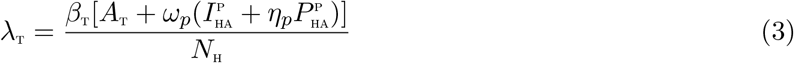

The parameters *κ*_t_(*κ*_t_ ≥ 1) is a modification parameter accounting for the increased infectiousness of dually infected individuals due to tuberculosis, *η*_t_(*η*_t_ ≤ 1) is a modification parameter accounting for reduced infectiousness of dually infected individuals due to latent TB. *ω*_*p*_(*ω*_*p*_ ≥ 1) is a modification parameter accounting for the increased infectiousness of dually infected individuals due to HPV while *η*_*p*_(*η*_*p*_ ≤ 1) is a modification parameter accounting for the reduced infectiousness of singly infected and dually infected individuals due to persistent HPV infection. This population is further reduced by natural death (at a rate *µ*_H_ natural death occurs in all epidemiological compartments at this rate). In (2), *β*_HP_ is the effective contact rate for transmission of HPV infection.

### 2.1 Basic properties of the co-infection model (1) without controls

The basic qualitative properties of the oncogenic HPV-TB co-infection model (1) will now be considered. Specifically, we establish the following results.

#### 2.1.1 Positivity and boundedness of solutions

For the oncogenic HPV-TB co-infection model (1) to be meaningful in the concept of epidemiological sense, it is necessary to prove that all its state variables are non-negative over time. The approach adopted by Omame *et al* [23] *will be used to establish the results below*.

##### Theorem 2.1

*Let the initial data be*
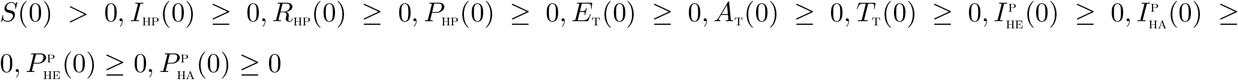

*Then the solutions*
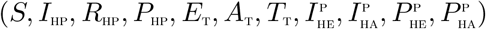 *of the model* (1) *are non-negative for all time t >* 0.

##### Lemma 2.1

*The region* 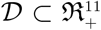 *is positively-invariant for the co-infection model* (1) *with initial conditions in* 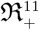.

### 3 Mathematical Analysis of the model without controls

#### 3.1 Basic reproduction number of the co-infection model

The co-infection model (1) has a DFE, obtained by setting the right-hand sides of the equations in the model (1) to zero, given by

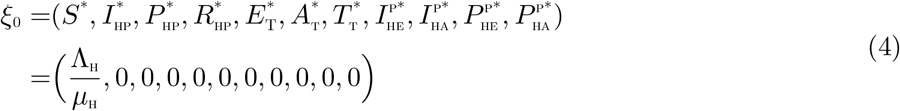

The basic reproduction number of the oncogenic HPV-TB co-infection model (1), using the approach in van den Driessche and Watmough [35], is given by ℛ_0_ = max{ℛ_0H_, ℛ_0T_} where ℛ_0H_ and ℛ_0T_ are, respectively, the oncogenic HPV and TB associated reproduction numbers, given by

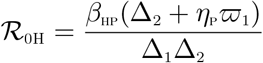

and

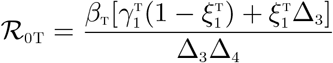

where,

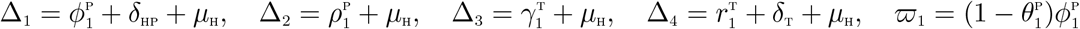

Using Theorem 2 of [35], the following result is established.

##### Lemma 3.1

*The DFE, ξ*_0_, *of the oncogenic HPV-TB co-infection model* (1) *is locally asymptotically stable if* ℛ _0_ *<* 1, *and unstable if* ℛ _0_ *>* 1.

### 3.2 Global asymptotic stability(GAS) of the disease-free equilibrium(DFE) *ξ*_0_ of the co-infection model

We shall apply the approach illustrated in [5] to investigate the global asymptotic stability of the disease free equilibrium of the co-infection model. In this section, we list two conditions that if met, also guarantee the global asymptotic stability of the disease-free state. First, system (1) must be written in the form:

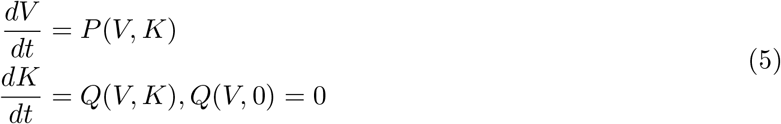

where *V* ∈ *R*^*m*^ denotes (its components) the number of uninfected individuals and *K* ∈ *R*^*n*^ denotes (its components) the number of infected individuals. *U*_0_ = (*V* ^*∗*^, 0) denotes the disease-free equilibrium of this system. The conditions (*W* 1) and (*W* 2) following must be satisfied in order to guarantee local asymptotic stability:

(*W* 1): For 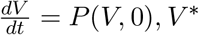 *V* ^*∗*^is globally asymptotically stable (GAS),

(*W* 2): 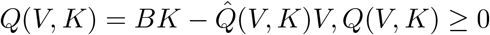 for (*V, K*) ∈ Ω,

where *B* = *D*_*K*_*Q*(*V* ^*∗*^, 0) is an M-matrix (the off-diagonal elements of *B* are nonnegative) and Ω is the region where the model makes biological sense. If System (1) satisfies the above two conditions then the following theorem holds:

#### Theorem 3.1

*The fixed point U*_0_ = (*V* ^*∗*^, 0) *is a globally asymptotic stable (GAS) equilibrium of* (1) *provided that R*_0_ *<* 1 *(LAS) and that assumptions* (*W* 1) *and* (*W* 2) *are met*

**Proof**

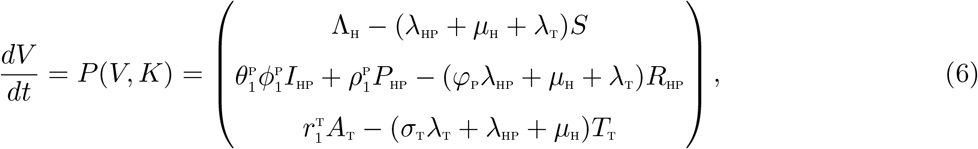

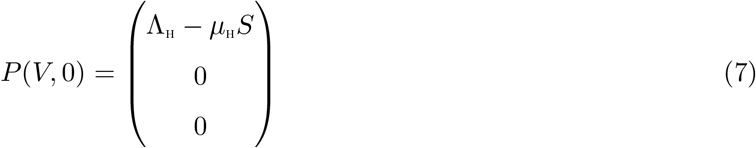

where *V* denotes the number of non-infectious compartments and *K* denotes the number of infectious compartments

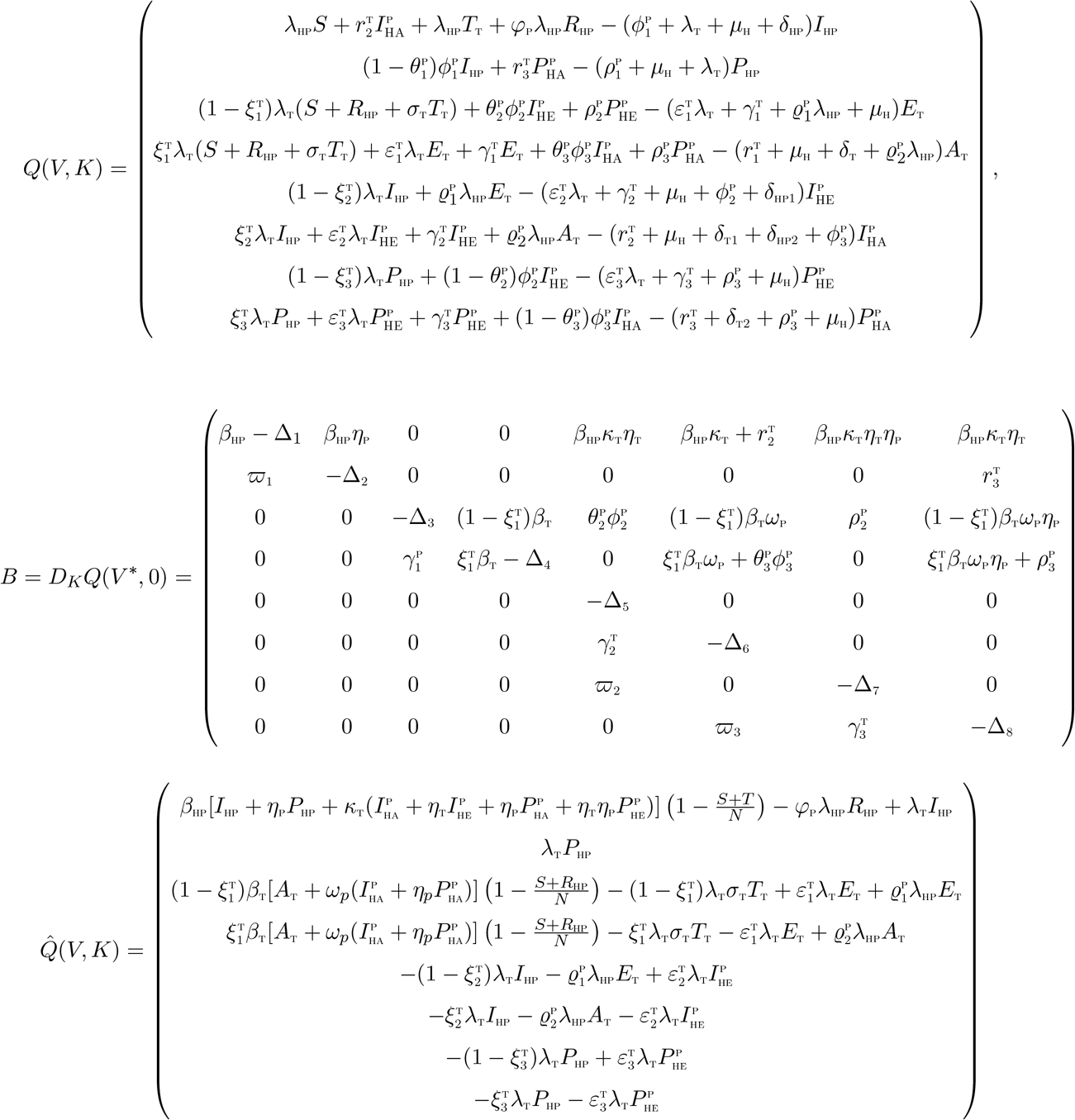

It is clear from the above, that, 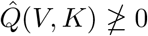. Hence the DFE may not be globally asymptotically stable, suggesting the possibility of a backward bifurcation. This supports the backward bifurcation analysis in the proceeding section.

### 3.3 Backward bifurcation analysis of the model without controls

We shall carry out analysis in this section to know the type of bifurcation the model (1) may undergo, using the Centre Manifold Theory as illustrated in [6]. The following result can be established.

#### Theorem 3.2

*Suppose a backward bifurcation coefficient a >* 0, *(with a defined below), when* ℛ_0_ < 1

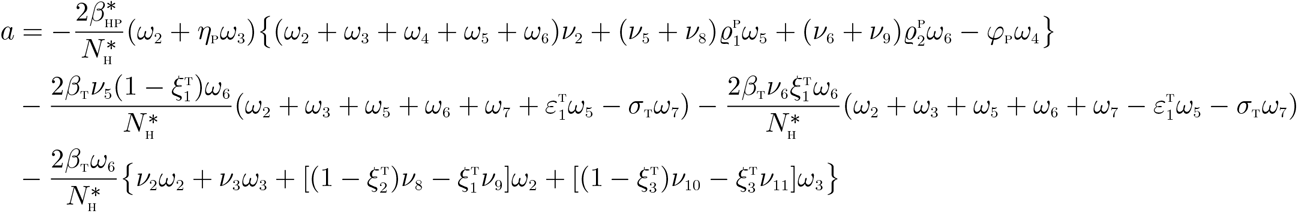

*then model* (1) *exhibits backward bifurcation at* ℛ_0_ = 1. *If a <* 0, *then the system* (1) *exhibits a forward bifurcation at* ℛ_0_ = 1.

**Proof**

Suppose

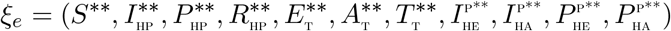

represents any arbitrary endemic equilibrium of the model. The existence of backward bifurcation will be studied using the Centre Manifold Theory [6]. To apply this theory, it is appropriate to do the following change of variables.

Let

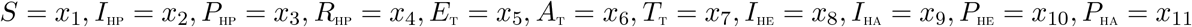

so that

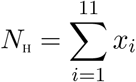

Further, using the vector notation

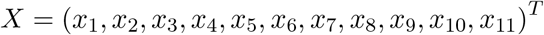

the model (1) can be re-written in the form

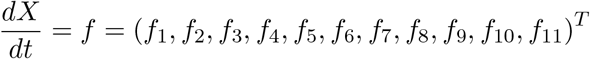

as follows:

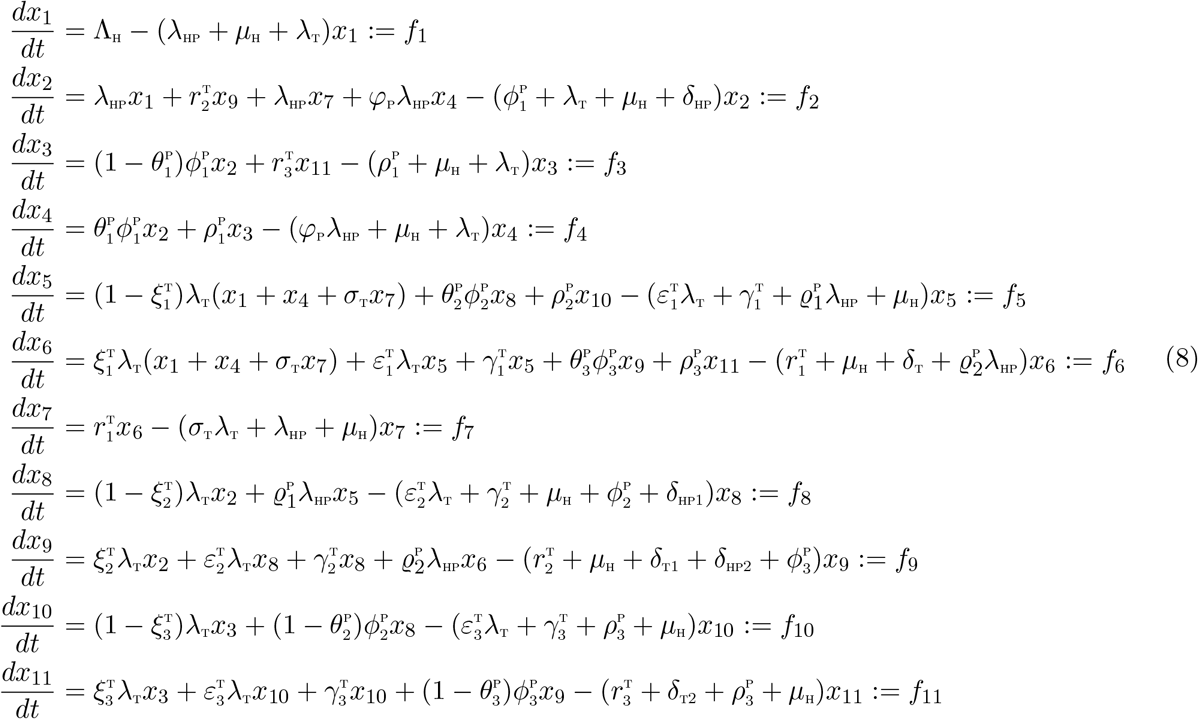

with:

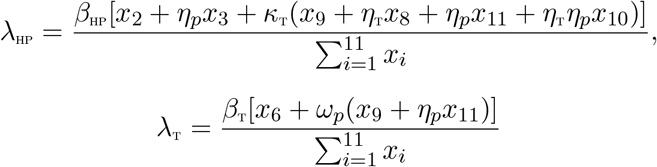

Consider the case when ℛ_0H_ = 1. Assume, further, that *β*_HP_ is chosen as a bifurcation parameter. Solving for 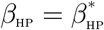 from ℛ_0H_ = 1 gives

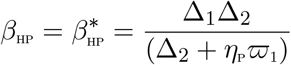

Evaluating the Jacobian of the system (8) at the DFE, *J*(*ξ*_0_), and using the approach in [6], we have that *J*(*ξ*_0_) has a right eigenvector (associated with the simple zero eigenvalue) given by

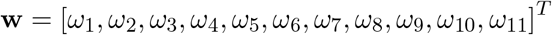

where,

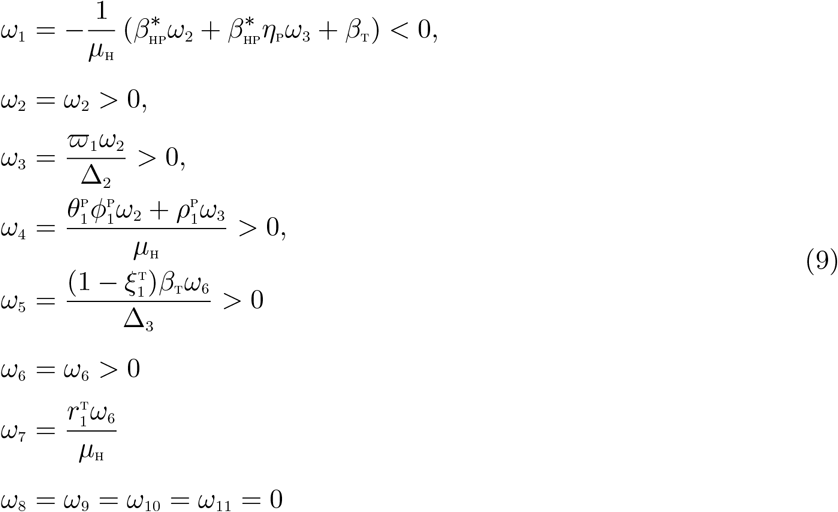

The components of the left eigenvector of 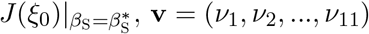, satisfying **v.w** = 1 are

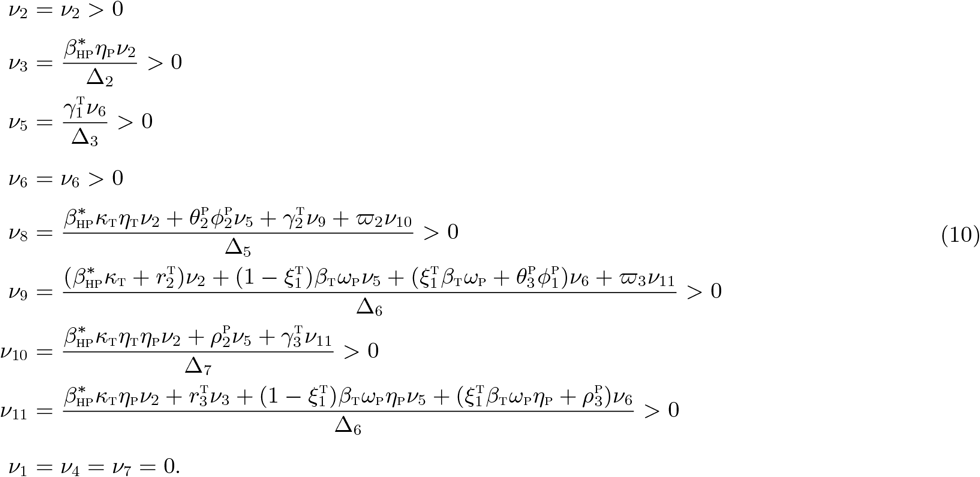

where,

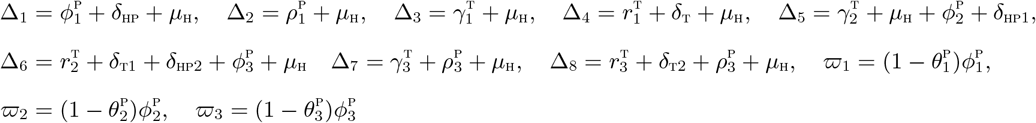

The associated bifurcation coefficients defined by *a* and *b*, given by:

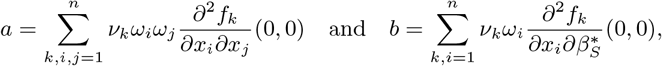

are computed to be

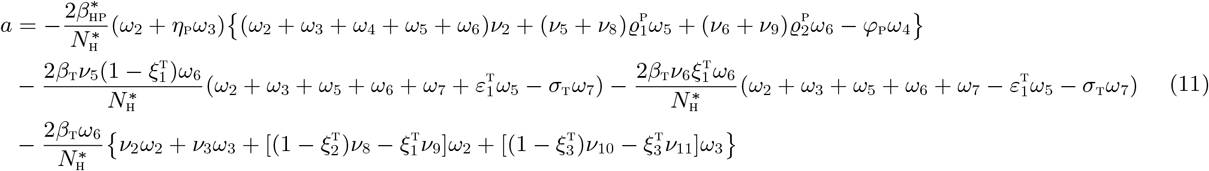

and

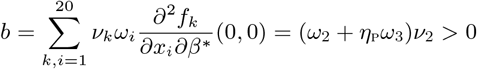

Since the bifurcation coefficient *b* is positive, it follows from Theorem 4.1 in [6] that the model (1), or the transformed model (8), will undergo a backward bifurcation if the backward bifurcation coefficient, *a*, given by (11) is positive. Setting the HPV and TB re-infection paramters *ϕ*_p_ = 0, *σ*_T_ = 0 and the TB exogenous re-infection term 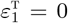, the bifurcation coefficient, 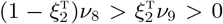 and 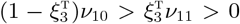, based on the definition of the components in (9), and also noting that all other components of the left and right eigenvectors occuring in the coefficient, *a*, are positive, including the parameters of the model. Hence, backward bifurcation does not occur in the Oncogenic HPV-TB co-infection model, in the absence of HPV and TB re-infection and in the absence of exogenous re-infection. □

## 4 Optimal control model

In this section, we shall use the Pontryagin ‘s Maximum Principle to determine the necessary conditions for the optimal control of the oncogenic HPV-TB co-infection model. We incorporate time dependent controls into the model (1) to determine the optimal strategy for curbing the co-infections of the two diseases. Thus, we have,

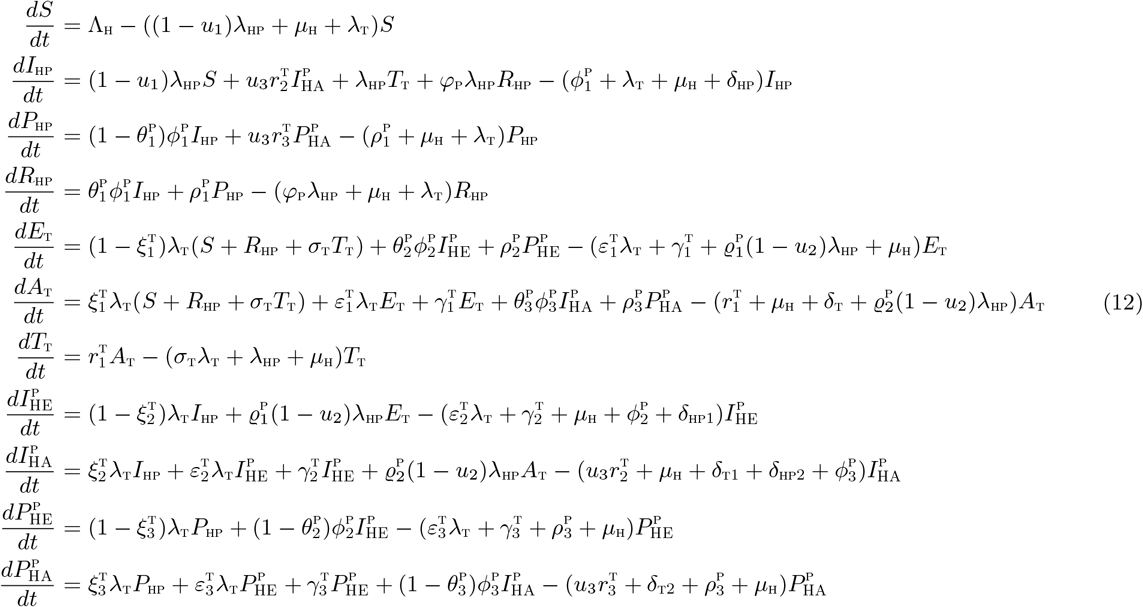

subject to the initial conditions 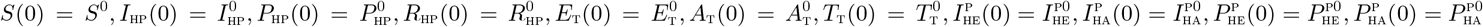

with:

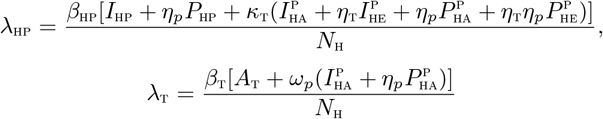

The control functions, *u*_1_(*t*), *u*_2_(*t*), and *u*_3_(*t*) are bounded, Lebesgue integrable functions. The control *u*_1_(*t*) represents the efforts at preventing incident HPV infections. The control *u*_2_(*t*) aims at preventing infection with HPV by TB infected individuals so as to reduce the co-infection cases. TB Treatment control for individuals dually infected with oncogenic HPV and TB is denoted by *u*_3_(*t*). The controls *u*_1_ and *u*_2_ satisfies 0 ≤ *u*_1_, *u*_2_ ≤ 1 while the control *u*_3_ satisfies 0 *< u*_3_ ≤ *h*, where *h* is the TB drug efficacy used for the treatment of co-infected individuals. Our optimal control problem involves a scenario where the number of HPV-infected, TB-infected, the co-infection cases and the cost of implementing preventive and treatment controls *u*_1_(*t*), *u*_2_(*t*) and *u*_3_(*t*) are minimized subject to the state system (12). For this, we consider the objective functional

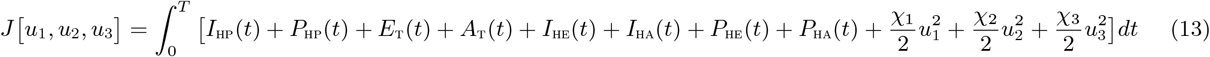

*T* is the final time. We seek to find an optimal control, 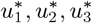, such that

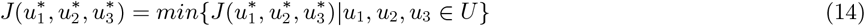

where 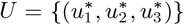 such that 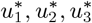 are measurable with 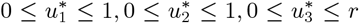, for *t* ∈ [0, *T*] is the control set. We shall now establish the existence of such an optimal solution which minimizes the objective functional *J*.

### Theorem 4.1

*Given the objective functional J, defined on the control set U, and subject to the state system* (12) *with non-negative initial conditions at t* = 0, *then there exists an optimal control triple u*^∗^ = (*u*_1_, *u*_2_, *u*_3_) *such that J*(*u*^∗^) = *min* {*J*(*u*_1_, *u*_2_, *u*_3_)|*u*_1_, *u*_2_, *u*_3_ ∈ *U*}.

We shall prove the existence of the given optimal control *u*^∗^, along with the corresponding solution trajectories by following the approach used in Mohammed-Awel and Numfor [18].

**Proof**

The state functions are positive and the controls are Lebesgue measurable, therefore we have that *J*(*u*_*i*_) ≥ 0 for all *u*_*i*_ ∈ *U*. As a result, 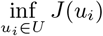 exists and is finite. Therefore, a minimizing sequnce of controls (*u*_*i*_) ∈ *U* exists such that

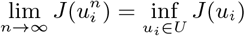

Let 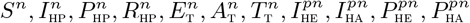 be the associated state trajectories. Since the state sequences are uniformly bounded, we have that the derivatives are also uniformly bounded. As a result, the state sequences are Lipschitz continuous with the same constant. Applying the Arzela-Ascoli Theorem [13], there exist 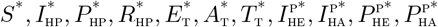 such that on a sub-sequence

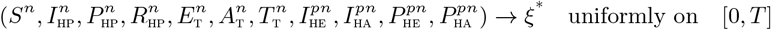

where,

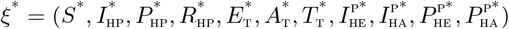

Since ‖*u*_*i*_‖_*L*_*∞ < K* for some *K >* 0, it follows that *u*_*i*_ ∈ *L*^2^ ([0, *T*]), such that on a sub-sequence

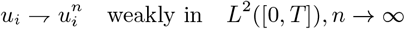

Applying the lower semi-continuity of *L*^2^ norm with respect to weak convergence, we have that

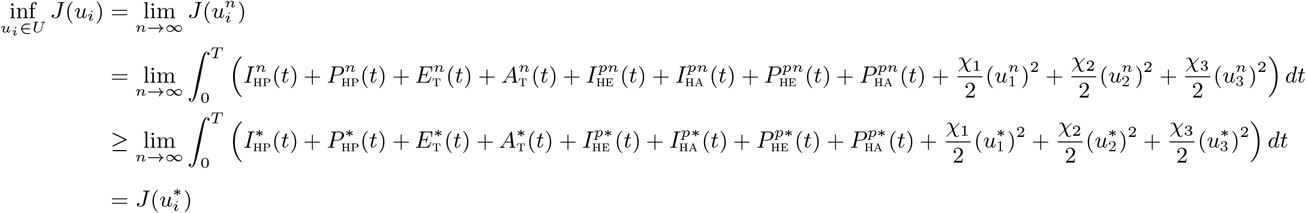

Considering the convergence of the sequences

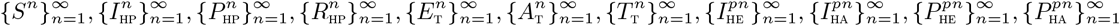

and passing to the limit in the ordinary differential equation system (12), we have that 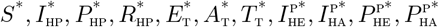 are the states corresponding to the control triple (*u*_*i*_). Hence, (*u*_*i*_) is an optimal control triple.

The Pontryagin ‘s Maximum Principle [28] gives the necessary conditions which an optimal control pair must satisfy. This principle transforms (12), (13) and (14) into a problem of minimizing a Hamiltonian, ℋ, pointwisely with regards to the control functions, *u*_1_, *u*_2_, *u*_3_:

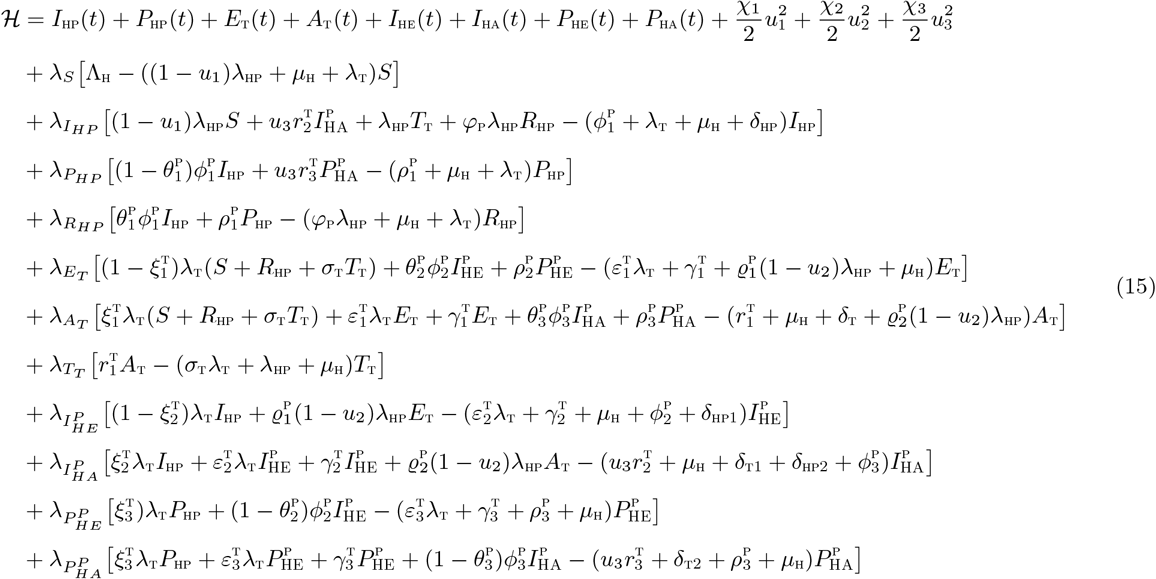

### Theorem 4.2

*For an optimal control set u*_1_, *u*_2_, *u*_3_ *that minimizes J over U, there are adjoint variables, λ*_1_, *λ*_2_, …, *λ*_11_ *Satisfying*

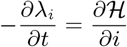

*and with transversality conditions*

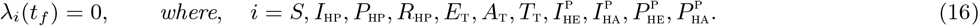

*Furthermore*,

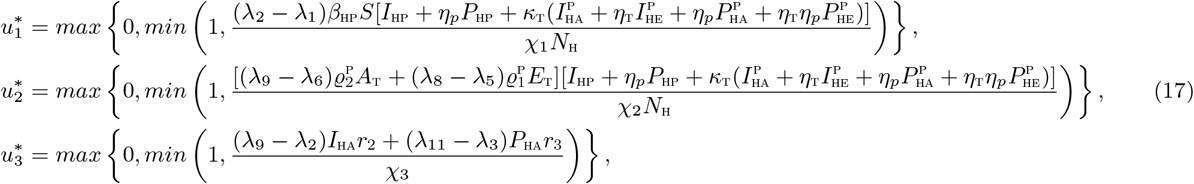

**Proof of Theorem 4.2**

Suppose 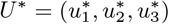 is an optimal control and 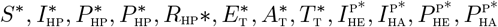 are the corresponding state solutions. Applying the Pontryagin ‘s Maximum Principle [28], there exist adjoint variables satisfying:

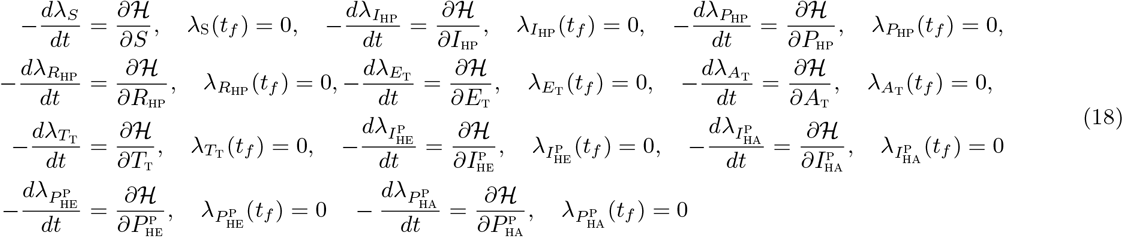

with transversality conditions;

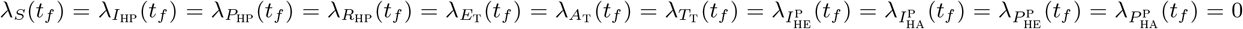

We can determine the behaviour of the control by differentiating the Hamiltonian, ℋ with respect to the controls(*u*_1_, *u*_2_, *u*_3_) at *t*. On the interior of the control set, where 0 *< u*_*j*_ *<* 1 for all (*j* = 1, 2, 3), we obtain

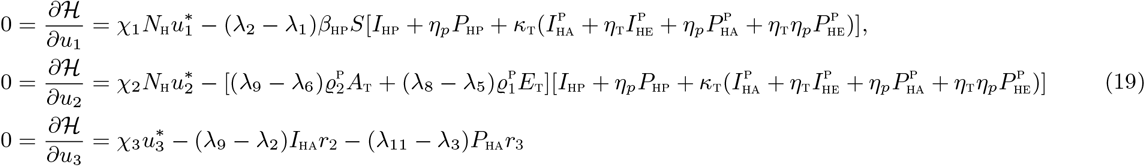

Therefore, we have that [14]

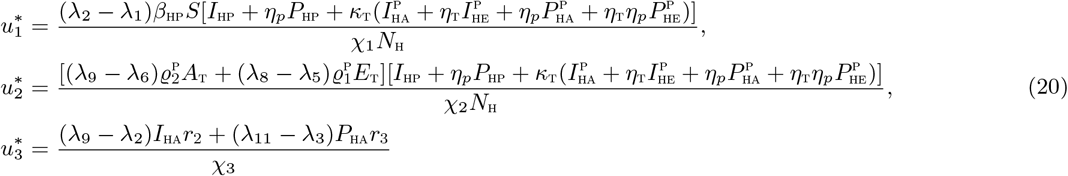

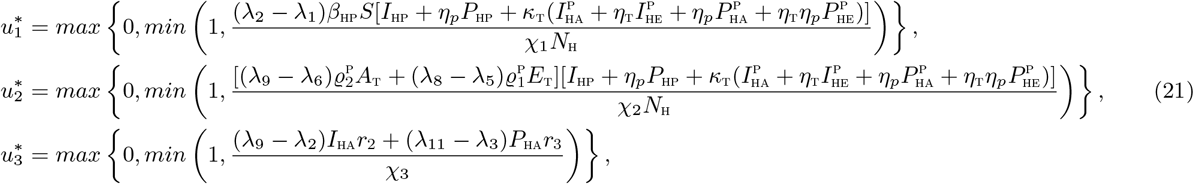

## 5 Simulations

In this section, we shall explore uncertainty and sensitivity analyses on the parameters of the model due to lack of precision in the estimates of some of the co-infection model parameters and the uncertainty which may occur when gathering data for the simulations. We shall also carry out numerical simulations of the optimal control model (12), in order to assess the impact of different intervention strategies on the dynamics of oncogenic HPV-TB co-infections.

### 5.1 Uncertainty and sensitivity analyses

The oncogenic HPV-TB co-infection model (1) has thirty nine (39) parameters, and uncertainties are expected to arise in the estimates of their values used in the numerical simulations. Adopting the approach used by Blower and Dowlatabadi [3], we shall carry out a Latin Hypercube Sampling (LHS) on the parameters of the model. For the sensitivity analysis, a Partial Rank Correlation Coefficient (PRCC) was calculated between values of the parameters in the response function and the values of the response function derived from the sensitivity analysis. A total of 1,000 simulations of the co-infection model (1) *per* LHS were run. Using the total number of individuals dually infected with HPV and latent 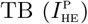 as the response function, the parameters that strongly influence the dynamics of the co-infection model (1) are the demographic parameter, *µ*_H_, the effective contact rate for HPV transmission, *β*_HP_, the parameter accounting for increased susceptibility to HPV by TB infected individuals, 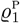, and the recovery rate from HPV for individuals in 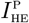 compartment. Also, using the population of individuals dually infected with HPV and active 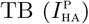 as the input, the five top ranked parameters are the effective contact rate for HPV transmissibility, *β*_HP_, the effective contact rate neccesary for TB transmission, *β*_T_, the modification parameter accounting for increased susceptibility to HPV infection by active TB infected individuals, 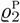, the recovery rate from HPV for individuals in 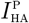compartment and the recovery rate from TB for individuals in 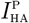 class. Taking the population of individuals dually infected with persistent HPV and latent 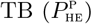 as the input, the three top ranked parameters are the effective contact rate for HPV transmissibility, *β*_HP_, the fraction of individuals who recover from HPV infection and do not progress to persistent HPV infection, 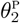 and the recovery rate from persistent HPV infection, 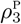, for individuals in 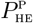 compartment. When the population of individuals dually infected with persistent HPV and active TB is used as the response function, the two top-ranked parameters are the effctive contact rate for TB transmission, *β*_T_ and the TB treatment rate, 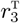, for individuals dually infected with persistent HPV and active TB.

Considering the HPV associated basic reproduction number, ℛ_0H_, as the response function, it is observed in Table 3 that the top four PRCC-ranked parameters are the demographic parameter, *µ*_H_, effective contact rate for HPV transmission, *β*_HP_, the fraction of infected individuals who recover from HPV and do not develop persistent HPV, 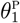 and the recovery rate from HPV 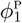. Finally, using the TB associated reproduction number, ℛ_0T_, as the response function the two key parameters that drive the dynamics of the model are the effective contact rate for TB transmission, *β*_t_ and the TB treatment ratein 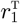.

**Table 3:** PRCC values for the co-infection model (1) parameters using the total number of individuals dually infected with: HPV and latent 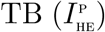 HPV and active 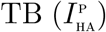, Persistent HPV and latent TB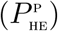, Persistent HPV and active 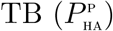, respectively, as well as the associated reproduction numbers *ℛ*_0H_ and *ℛ*_0T_, as response functions. Paramters which strongly inuence the dynamics of the co-infection model with respect to each of the response functions are shown in bold fonts.

| Parameters | $I_{\text{HE}}^p$ | $I_{\text{HA}}^p$ | $P_{\text{HE}}^p$ | $P_{\text{HA}}^p$ | $\mathcal{R}_{0\text{H}}$ | $\mathcal{R}_{0\text{T}}$ |
| --- | --- | --- | --- | --- | --- | --- |
| $\mu_{\text{H}}$ | **-0.4468** | -0.1478 | -0.3209 | 0.1208 | **-0.5074** | -0.2068 |
| $\beta_{\text{HP}}$ | **0.9304** | **0.6982** | **0.6390** | 0.3300 | **0.9165** | — |
| $\beta_{\text{T}}$ | 0.3353 | **0.5813** | -0.0719 | **0.5620** | — | **0.8182** |
| $\varrho_1^p$ | **0.5387** | 0.0493 | 0.1298 | 0.0116 | — | — |
| $\varrho_2^p$ | 0.1403 | **0.5009** | 0.0683 | 0.1199 | — | — |
| $\kappa_{\text{T}}$ | 0.1835 | 0.2371 | 0.0167 | 0.0904 | — | — |
| $\theta_1^p$ | -0.3717 | -0.0286 | -0.1216 | -0.0579 | **-0.7015** | — |
| $\theta_2^p$ | 0.1408 | 0.0140 | **-0.7032** | -0.0782 | — | — |
| $\theta_3^p$ | 0.0097 | 0.0652 | 0.0233 | -0.2433 | — | — |
| $\phi_1^p$ | -0.1802 | -0.0323 | -0.0447 | -0.0534 | **-0.8041** | — |
| $\phi_2^p$ | **-0.6866** | **-0.4079** | -0.2726 | -0.0732 | — | — |
| $\phi_3^p$ | 0.0394 | -0.2870 | -0.0175 | 0.2939 | — | — |
| $r_1^{\text{T}}$ | -0.0480 | -0.0471 | 0.000319 | 0.1303 | — | **-0.5629** |
| $r_2^{\text{T}}$ | 0.0805 | **-0.8312** | 0.2174 | 0.1088 | — | — |
| $r_3^{\text{T}}$ | 0.0477 | 0.1487 | 0.3076 | **-0.8994** | — | — |
| $\rho_1^p$ | -0.0891 | 0.0035 | -0.0376 | -0.0340 | — | — |
| $\rho_2^p$ | 0.1337 | 0.0170 | 0.0631 | 0.0208 | — | — |
| $\rho_3^p$ | -0.0734 | -0.0162 | **-0.5220** | -0.3034 | — | — |
| $\varphi_{\text{P}}$ | 0.0443 | -0.0039 | 0.0061 | -0.0184 | — | — |
| $\delta_{\text{HP}}$ | 0.0120 | 0.0144 | 0.0208 | 0.0337 | -0.0478 | — |
| $\delta_{\text{HP1}}$ | 0.0220 | -0.0026 | -0.0104 | 0.0085 | — | — |
| $\delta_{\text{HP2}}$ | -0.0418 | -0.0033 | -0.0214 | 0.0213 | — | — |
| $\delta_{\text{T}}$ | 0.0275 | -0.0132 | 0.0270 | -0.0041 | — | -0.1606 |
| $\delta_{\text{T1}}$ | 0.0016 | -0.1478 | 0.0101 | 0.0232 | — | — |
| $\delta_{\text{T2}}$ | 0.0097 | 0.0232 | 0.0324 | -0.1941 | — | — |
| $\sigma_{\text{T}}$ | 0.0062 | -0.0006321 | 0.0229 | -0.00886 | — | — |
| $\xi_1^{\text{T}}$ | -0.0687 | 0.0941 | -0.0301 | 0.0407 | — | 0.3606 |
| $\xi_2^{\text{T}}$ | 0.0377 | 0.0611 | 0.3845 | 0.0232 | — | — |
| $\xi_3^{\text{T}}$ | 0.0077 | -0.0035 | -0.0143 | 0.0111 | — | — |
| $\varepsilon_1^{\text{T}}$ | -0.0116 | 0.0231 | -0.0132 | -0.0138 | — | — |
| $\varepsilon_2^{\text{T}}$ | 0.0052 | 0.0445 | 0.0188 | 0.047 | — | — |
| $\varepsilon_3^{\text{T}}$ | 0.0058 | 0.0202 | -0.0116 | 0.0927 | — | — |
| $\gamma_1^{\text{T}}$ | -0.0297 | 0.0089 | 0.049 | 0.0153 | — | 0.0793 |
| $\gamma_2^{\text{T}}$ | -0.0237 | 0.0842 | -0.0314 | -0.0003668 | — | — |
| $\gamma_3^{\text{T}}$ | -0.0127 | -0.0199 | -0.0326 | 0.0583 | — | — |
| $\eta_{\text{T}}$ | 0.0731 | 0.1261 | 0.0344 | 0.0318 | — | — |
| $\eta_{\text{P}}$ | 0.1506 | 0.1929 | 0.0470 | 0.1196 | — | — |
| $\omega_{\text{P}}$ | 0.0475 | 0.1403 | -0.0277 | 0.1404 | — | — |

### 5.2 Numerical simulations

Numerical simulations of the optimal control problem (12), adjoint equations (18) and characterizations of the control (21) are implemented by the Runge Kutta method using the forward backward sweep (carried out in MATLAB). Following the choice of the weight function for HPV vaccination control used in Malik *et al* [17], *the balancing factor χ*_1_ = 10^3^. We also assume *χ*_2_ = 500 and *χ*_3_ = 400. The demographic data related to the Shanxi province in rural China is used [8]. We assume the initial conditions to be: 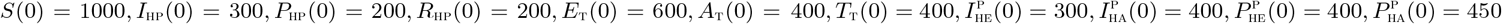,.We implement the following three different control strategies for numerical simulations of the co-infection model (4). **The unit of time is years, throughout the entire simulations**.

a. Strategy A: Control against incident HPV infection (*u*_1 ≠_0) and control against infection with HPV by TB infected individuals(*u*_2_ ≠ 0);
b. Strategy B: Control against incident HPV infection (*u*_1_ ≠ 0) and TB treatment control for dually infected individuals (*u*_3_ ≠ 0);
c. Strategy C: control against infection with HPV by TB infected individuals (*u*_2_ ≠ 0) and TB treatment control for dually infected individuals (*u*_3_ ≠ 0).

#### 5.2.1 Strategy A: Control against incident HPV infection (*u*_1_ ≠ 0) and control against Cinfection with HPV by TB infected individuals (*u*_2_ ≠ 0)

Applying this control strategy, we note that the total number of individuals dually infected with HPV and TB in latent and active stages, respectively, (Figures 2 (a) and 2(b)), and the total number of individuals dually infected with persistent HPV and TB in latent and active stages, respectively (Figures 2(c) and 2(d)) are less when this control is applied than when the control is not applied. To be specific, this control strategy averts almost 420,352 new co-infection cases, which is the greatest number of averted cases in comparison with other control strategies implemented. The control profile for this strategy given in Figure 5, reveals that control *u*_1_ is at its upper bound for the first 3.9 years before ultimately declining to zero. Similarly, the control *u*_2_ is at the maximum value of 90% for the first 8 months before declining to zero at time, *t*≠4.5 years.

#### 5.2.2 Strategy B: Control against incident HPV infection (*u*_1_ ≠ 0) and TB treatment control for dually infected individuals (*u*_3_ ≠ 0)

The simulations of the total number of dually infected individuals in the presence of TB treatment controls are depicted in Figures 3(a)-3(d). Applying this control, we observe that the total number of individuals dually infected with HPV and TB in latent and active stages of infection respectively, is less than the total population when no control is applied as expected (Figure 3(a) and 3(b)). Similarly, it is noticed from Figures (3(c) and 3(d), that high population level impact is observed in the total number of individuals dually infected with persistent HPV and TB in latent and active stages of infection, respectively, when this control strategy is applied. Specifically, when this control strategy is implemented, about 300,549 new co-infection cases were averted. The control profile for this strategy presented in Figure 7 shows that control *u*_1_ is at its upper bound for the first 4.0 years before steadily declining to zero at final time. Also, the control *u*_3_ is at the maximum value of 100% for the first 1.2 years and then steadily declines to zero at time, *t*≠4.5 years. This simulation results conforms with the epidemiological report in the introduction section that gynaecologic TB is a risk factor for oncogenic HPV infection (and subsequent cancer infection) [37]. Hence, if we focus on TB treatment controls, it can significantly reduce the burden of the co-infection of oncogenic HPV and TB in a population. The simulations are also in line with the point opined by [37], that prior TB infection was associated with persistent HPV and increased susceptibility to cervical cancer. As a result, treating TB infections in dually infected individuals will significantly curb the mixed infections of both diseases.

**Figure 2:**
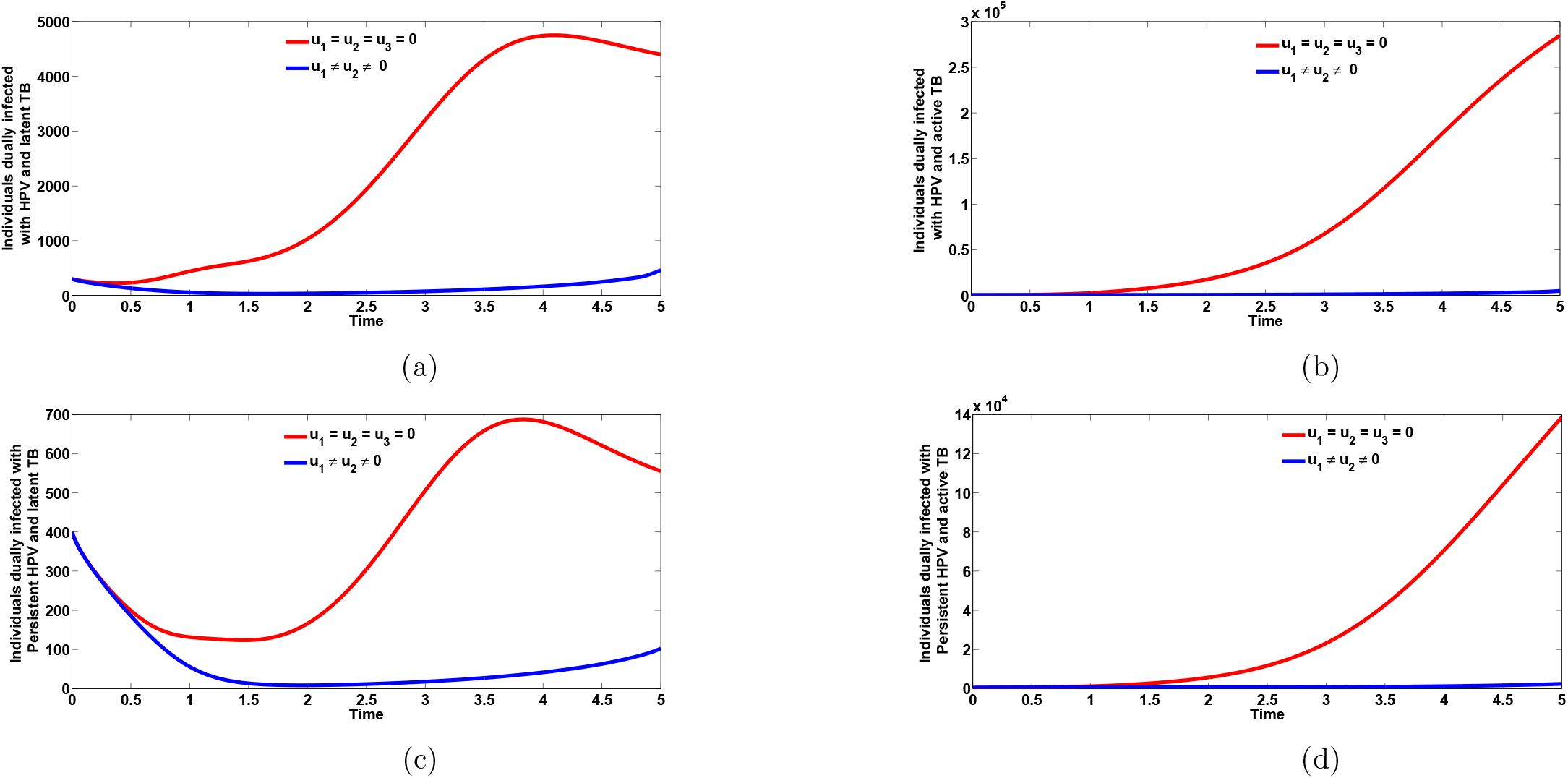
Plots of the total number of individuals dually infected with HPV and TB in latent and active stages, respectively (Figures 2a and 2b) as well as total number of individuals dually infected with persistent HPV and TB in latent and active stages (Figures 2c and 2d), respectively, the presence of optimal vaccination control against incident HPV infection (*u*_1_ ≠ 0) and control against infection with HPV by TB infected individuals (*u*_2_ ≠ 0). Here, *β*_HP_ ≠ 2.0, *β*_T_ ≠ 8.557. All other parameters as in Table 2

**Figure 3:**
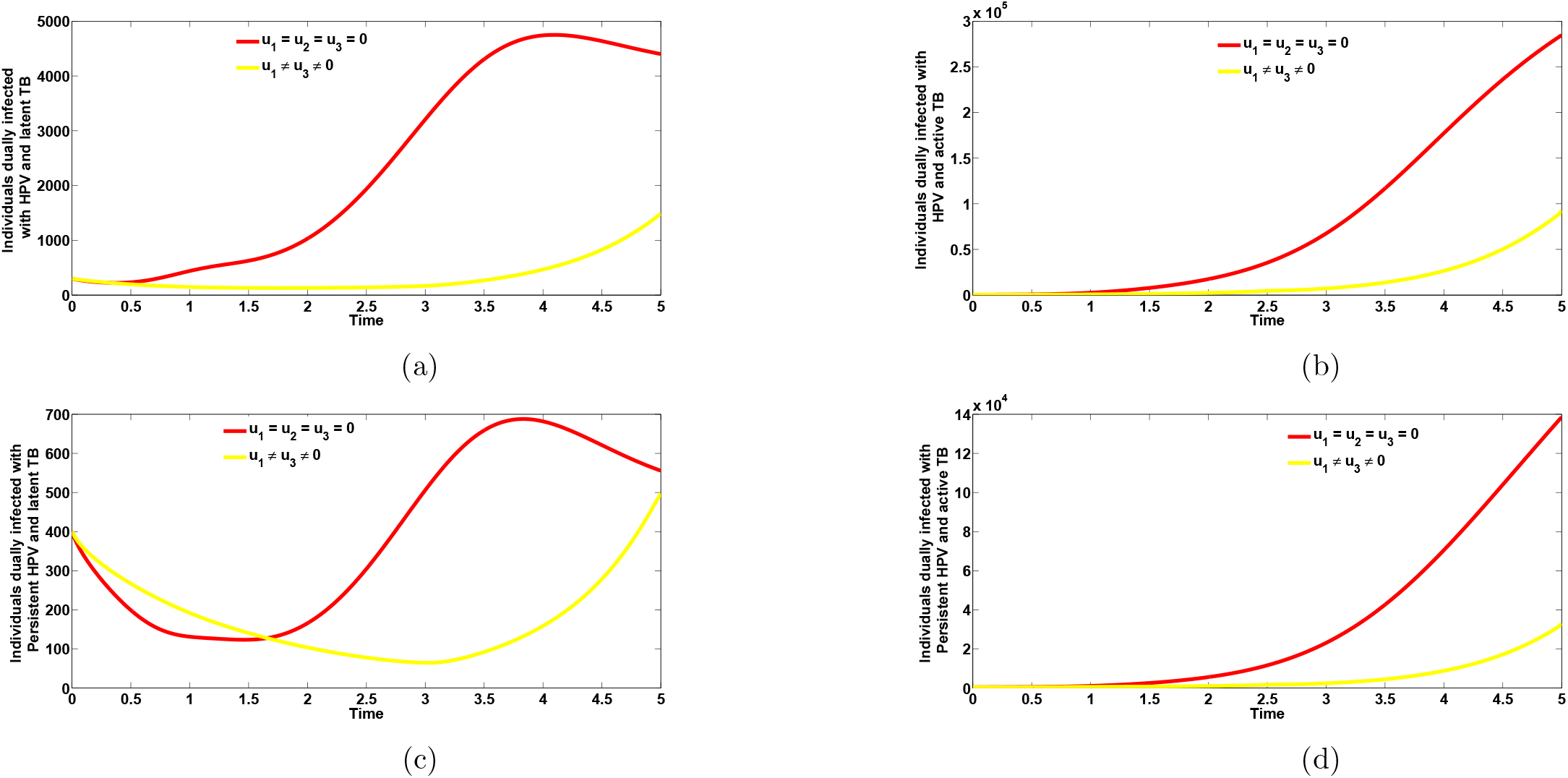
Plots of the total number of individuals dually infected with HPV and TB in latent and active stages, respectively (Figures 3(a) and 3(b) as well as total number of individuals dually infected with persistent HPV and TB in latent and active stages (Figures 3(c) and 3(d), respectively, the presence of optimal vaccination control against incident HPV infection (*u*_1_ = 0) and TB treatment control dually infected individuals (*u*_3_ = 0). Here, *β*_HP_ = 2.0, *β*_T_ = 8.557. All other parameters as in Table 2

**Figure 5:**
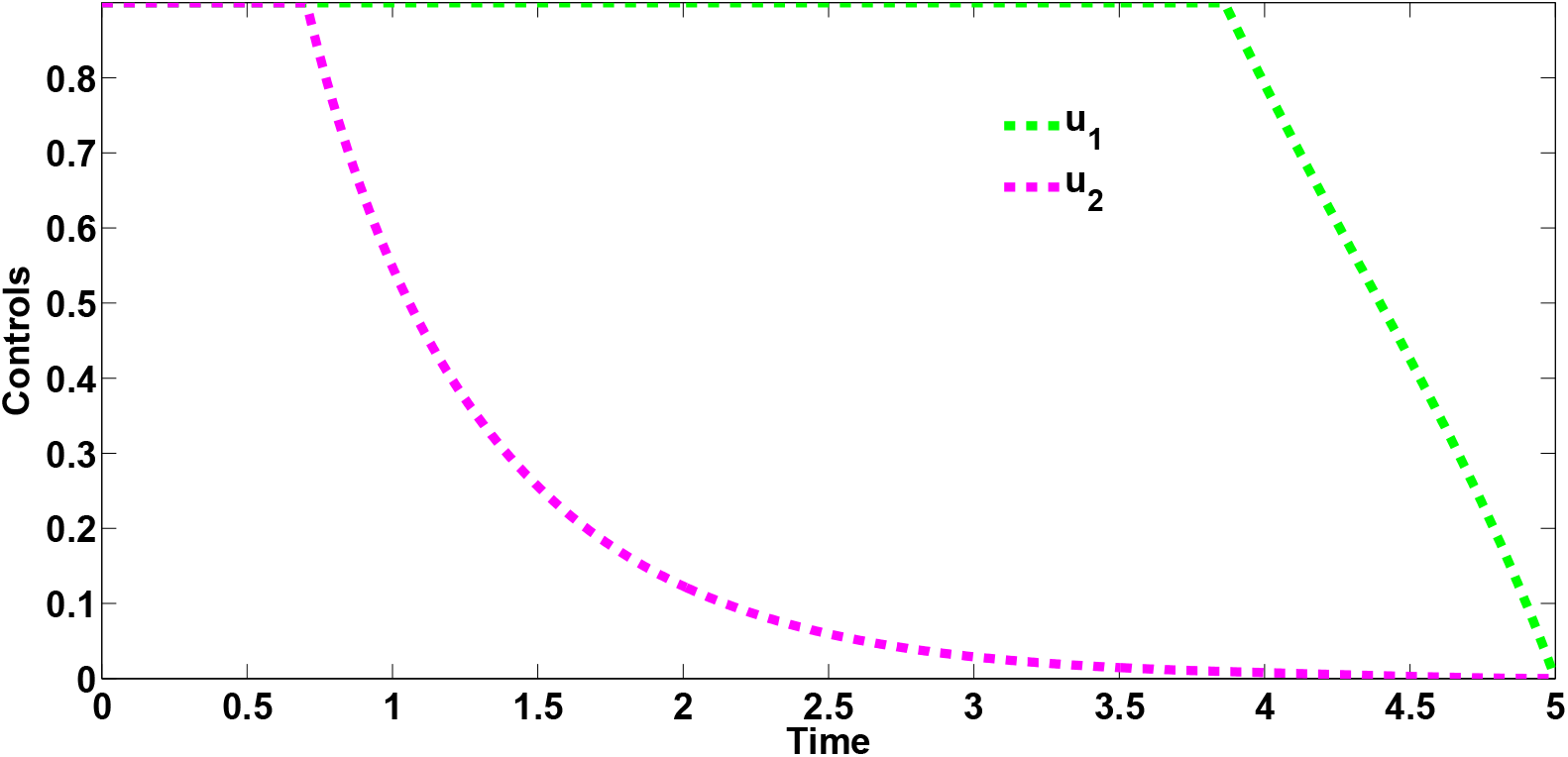
Combined effects of optimal controls *u*_1_ and *u*_2_ on the dynamics of the co-infection optimal control model (12)

**Figure 7:**
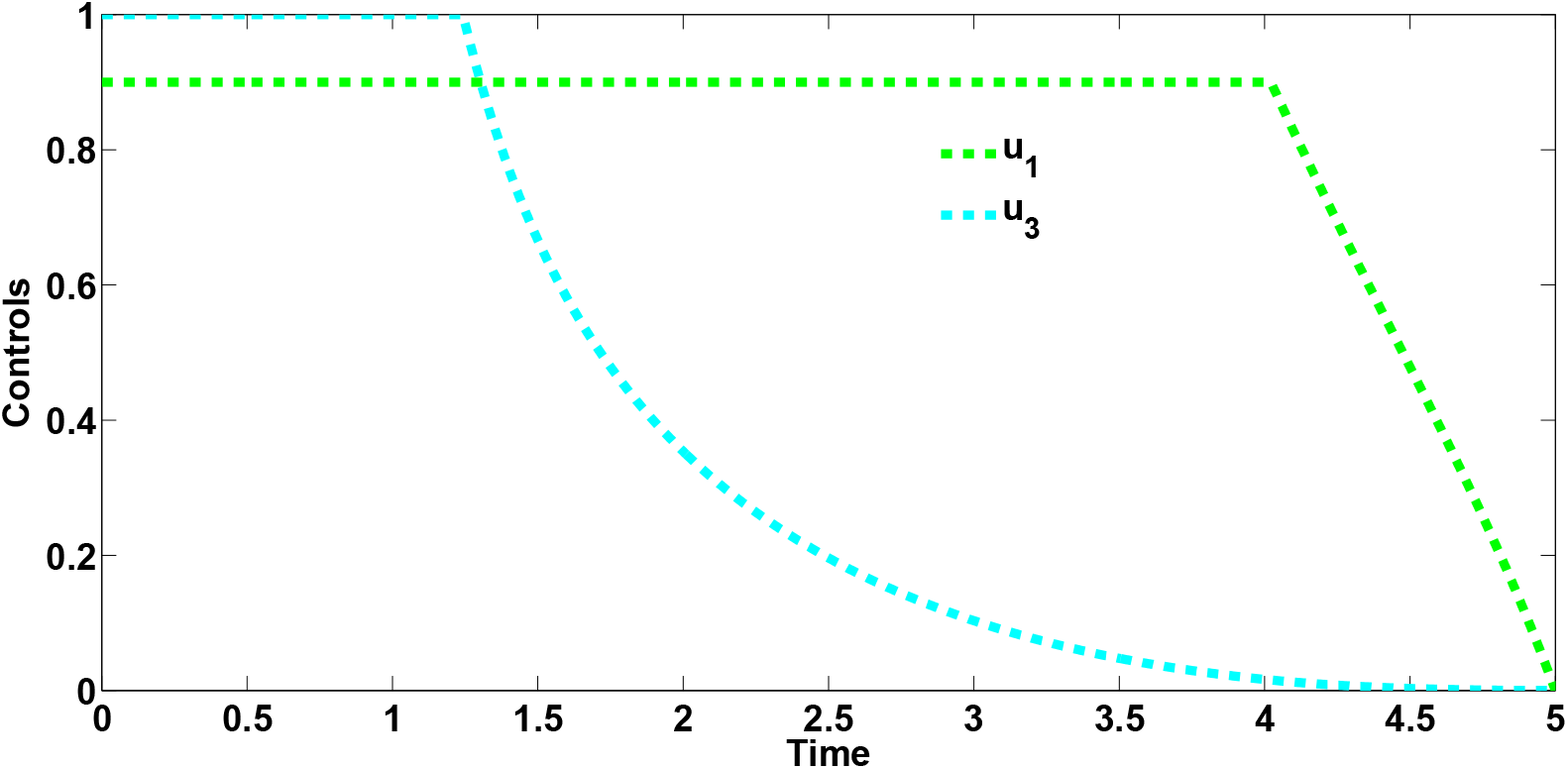
Combined effects of optimal controls *u*_1_ and *u*_3_ on the dynamics of the co-infection optimal control model (12)

#### 5.2.3 Strategy C: control against infection with HPV by TB infected individuals (*u*_2_ ≠ 0) and TB treatment control for dually infected individuals (*u*_3_ ≠ 0)

Using this control strategy, we observe in Figures 10(a) and 10 (b), that the number of individuals dually infected with HPV and TB in latent and active stages of infection, respectively, is less than the number when no control strategy is applied. Likewise, this strategy has positive population level impact on the populations of infected individuals dually infected with persistent HPV and TB in latent and active stages of infection, respectively, Figures 10(c) and 10 (d). When this intervention strategy is administered, approximately 362,573 new co-infection cases are averted. The control profile for this strategy, depicted by Figure 9 reveals that control *u*_2_ is at its upper bound for the first 4.5 years before gradually falling down to zero at final time. Moreso, the control *u*_3_ is at its minimum value for the first 2.1 years and then steadily rises to its peak value of 100% at time, *t*=2.7 years, before gradually declining to zero at final time. It is imperative to state categorically, that the results of the simulations are based on the parameter values given in table 2 and the initial conditions and weight constants given in Section 5.2

**Figure 9:**
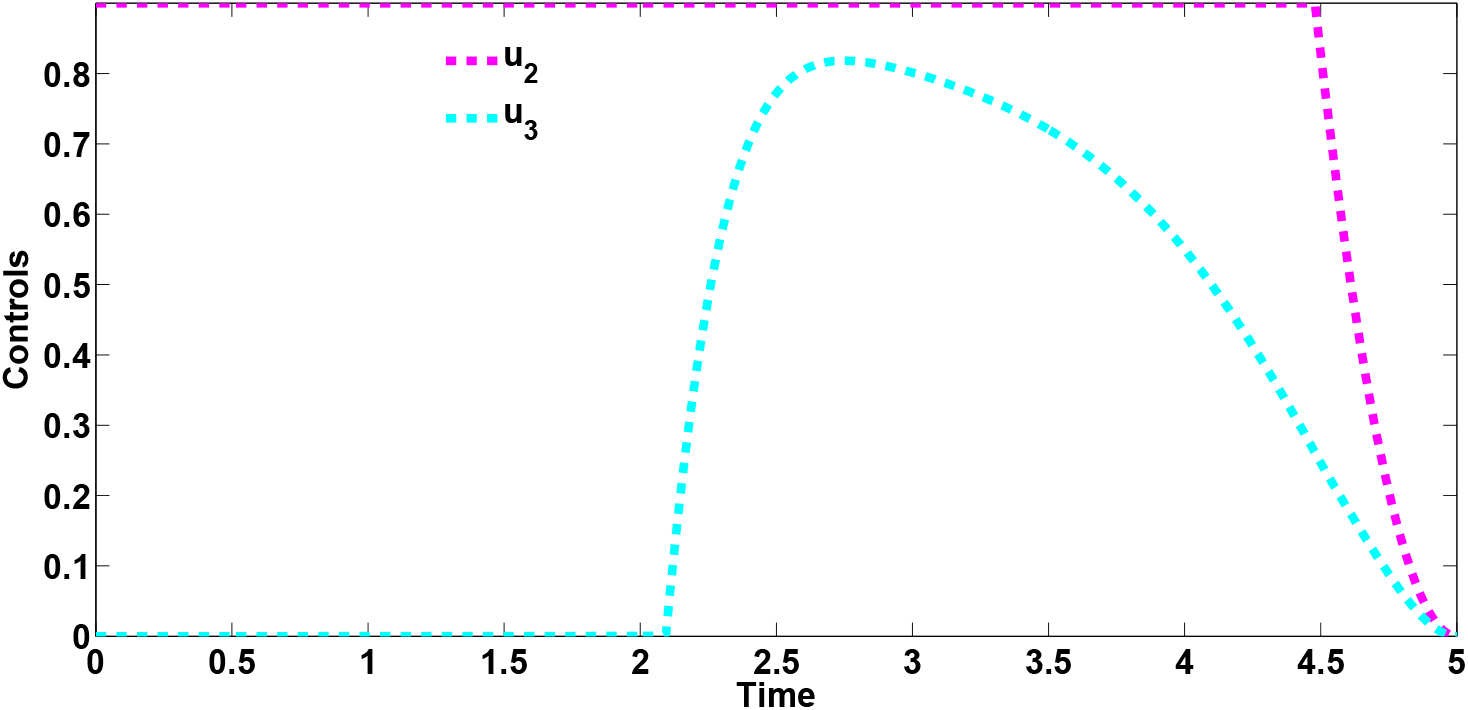
Combined effects of optimal controls *u*_2_ and *u*_3_ on the dynamics of the co-infection optimal control model (12)

**Figure 10:**
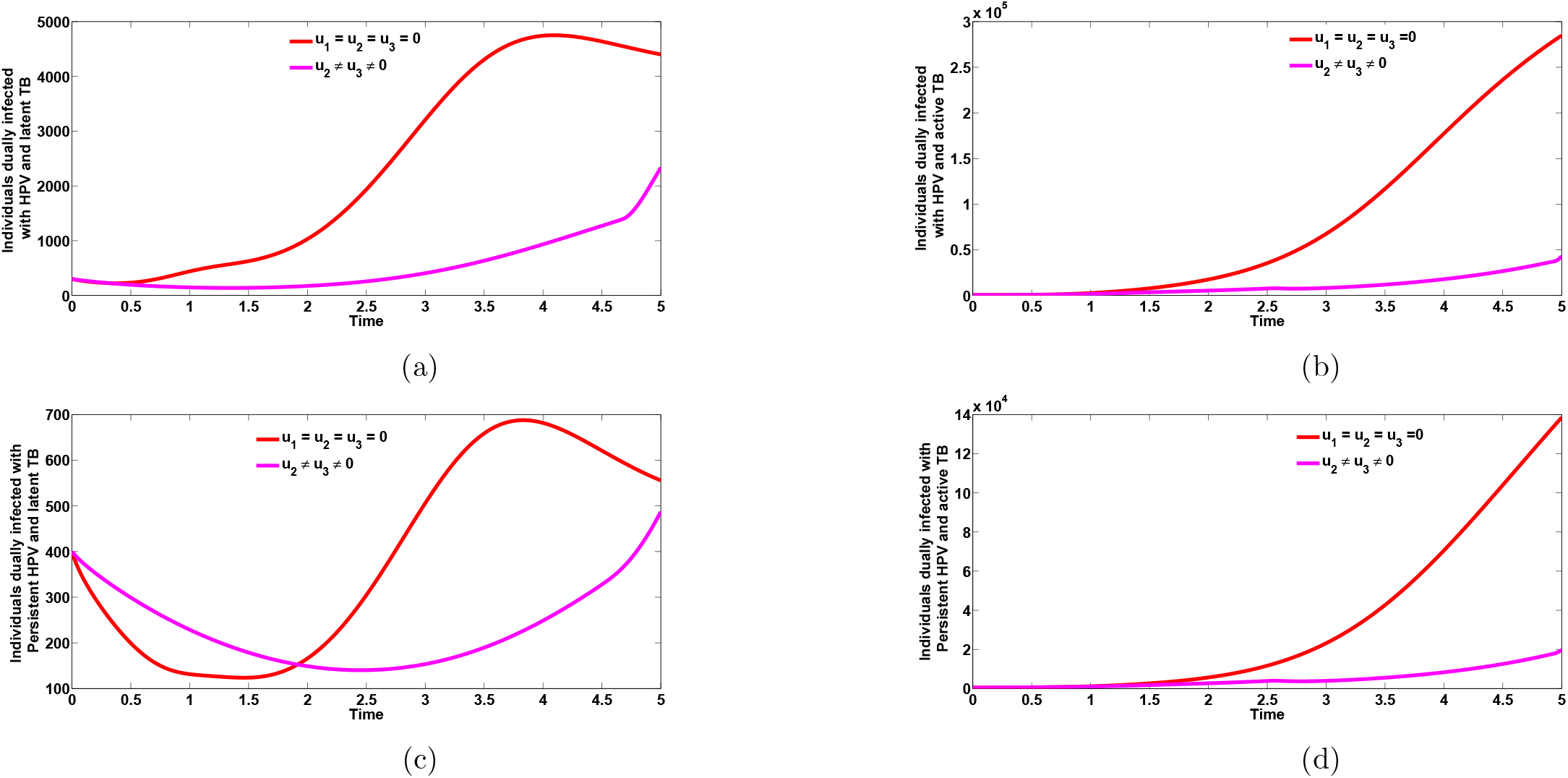
Plots of the total number of individuals dually infected with HPV and TB in latent and active stages, respectively (Figures 10a and 10b) as well as total number of individuals dually infected with persistent HPV and TB in latent and active stages (Figures 10c and 10d), respectively, in the presence of control against HPV infection by TB-infected individuals (*u*_2_ ≠ 0) and TB treatment control dually infected individuals (*u*_3_ ≠ 0). Here, *β*_HP_ ≠ 2.0, *β*_T_ ≠ 8.557. All other parameters as in Table 2

**Figure 12:**
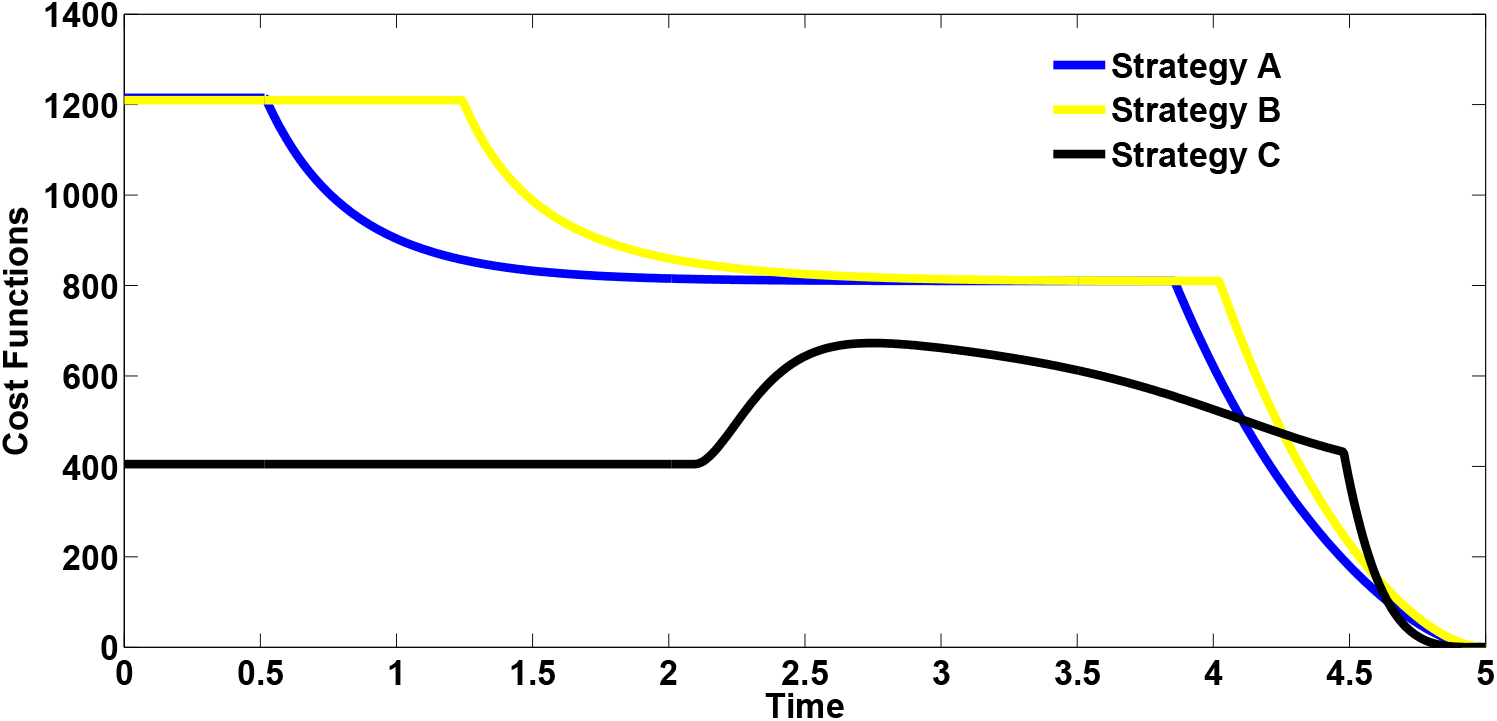
Cost functions of the different control strategies

### 5.3 Cost-effectiveness analysis

The cost-effectiveness analysis is used to evaluate the health interventions related benefits so as to justify the costs of the strategies [4]. This is obtained by comparing the differences among the health outcomes and costs of those interventions; achieved by computing the incremental cost-effectiveness ratio (ICER), which is defined as the cost per health outcome. It is given by:

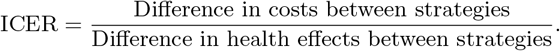

We calculated the total number of co-infection cases averted and the total cost of the strategies applied in Table 4. The total number of co-infection cases prevented is obtained by calculating the total number of individuals when controls are administered and the total number when control is not applied. Similarly, we apply the cost functions 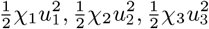, over time, to compute the total cost for the various strategies that we implemented. We now compare the cost-effectiveness of strategy B (Optimal HPV vaccination control for sexually active susceptible individuals against incident HPV infection and TB treatment control for dually infected individuals) and strategy C (Control against HPV infection by TB infected individuals and TB treatment controls for dually infected individuals).

**Table 4:** Increasing order of the total infection averted due to the control strategies

| Strategy | Total infection averted | Total cost | ACER | ICER |
| --- | --- | --- | --- | --- |
| B: $u_1(t), u_2(t)$ | 300, 549.2 | 1, 210 | 0.004026 | 0.004026 |
| C: $u_2(t), u_3(t)$ | 362, 573.4 | 405 | 0.001117 | -0.01298 |
| A: $u_1(t), u_2(t)$ | 420, 352.2 | 1, 215 | 0.002890 | 0.01402 |

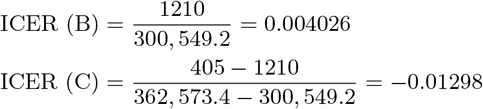

From ICER (B) and ICER(C), we observe a cost saving of 0.01298 observed for strategy C over strategy B. This implies that strategy B strongly dominated strategy C, showing that strategy B is more costly and less effective compared to strategy C. Therefore, strategy B is removed from subsequent ICER computations, shown in Table 5. We shall now compare strategies C and A. Comparing strategy C (Control against HPV infection by TB infected individuals and TB treatment controls for dually infected individuals) and strategy A (Optimal HPV vaccination control for sexually active susceptible individuals against incident HPV infection and Control against HPV infection by TB infected individuals), we observe that ICER (A) is greater than ICER (C), showing that strategy A strongly dominated strategy C and is more expensive and less effective compared to strategy C. Consequently, strategy C (the strategy that combines and implements control against HPV infection by TB infected individuals as well as TB treatment controls for dually infected individuals) has the least ICER and is the most cost-effective of all the control strategies for the control and management of oncogenic HPV and TB co-infection. This clearly agrees with the results obtained from ACER method in Table 4 that strategy C is the most cost-effective strategy.

**Table 5:** Increasing order of the total infection averted due to the control strategies

| Strategy | Total infection averted | Total cost | ACER | ICER |
| --- | --- | --- | --- | --- |
| C: $u_2(t), u_3(t)$ | 362, 573.4 | 405 | 0.001117 | 0.001117 |
| A: $u_1(t), u_2(t)$ | 420, 352.2 | 1, 215 | 0.002890 | 0.01402 |

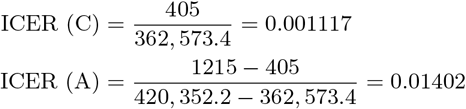

## 6 Conclusion

In this work, we have developed and presented a co-infection model for Oncogenic HPV and TB with cost-effectiveness optimal control analysis. The full co-infection model was shown to undergo the phenomenon of backward bifurcation when the associated reproduction number is less than unity. It was further shown that TB and HPV re-infection parameters (*ϕ*_p_ ≠ 0 and *σ*_t_ ≠ 0) as well as the exogenous re-infection term (*ε*_1_ ≠ 0) induced the phenomenon of backward bifurcation _1he_ in the oncogenic HPV-TB co-infection model. The global asymptotic stability of the disease-free equilibrium of the co-infection model was also proven **not to exist**, when the associated reproduction number was below unity. The necessary conditions for the existence of optimal control and the optimality system for the co-infection model was established using the Pontryagin ‘s Maximum Principle. Uncertainty and global sensitivity analyses were also carried out to determine the top ranked parameters that influence the dynamics of the co-infection model, when the associated reproduction numbers as well as the infected populations were used as response functions. When the population of individuals dually infected with HPV and latent 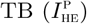 was used as the response function, the parameters that strongly influence the dynamics of the co-infection model (1) are the demographic parameter, *µ*_H_, the effective contact rate for HPV transmission, *β*_HP_, the parameter accounting for increased susceptibility to HPV by TB infected individuals, 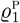, and the recovery rate from HPV hefor individuals in 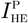 compartment. In addition, using the population of individuals dually infected with HPV and active 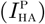 as the input, the five top ranked parameters are the effective contact rate for HPV transmissibility, *β*_HP_, the effective contact rate neccesary for TB transmission, *β*_T_, the modification parameter accounting for increased susceptibility to HPV infection by active TB infected individuals, 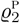, the recovery rate from HPV for individuals in 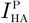 compartment and the recovery rate from TB for individuals in 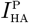 class. Taking the population of individuals dually infected with persistent HPV and active TB as the response function, the two top-ranked parameters are the effctive contact rate for TB transmission, *β*_*T*_and the TB treatment rate, 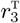, for individuals dually infected with persistent HPV and active TB. Numerical simulations of the optimal control model showed that:

i. A combination of HPV prevention control and TB treatment control has a positive population level impact in reducing the burden of oncogenic HPV and TB co-infection cases in a population.
ii. A strategy that implements control against HPV infection by TB infected individuals and TB treatment controls for dually infected individuals can significantly reduce the burden of oncogenic HPV and TB co-infections.
iii. The strategy that combines and implements control against HPV infection by TB infected individuals as well as TB treatment control for dually infected individuals has the least ICER and is the most cost-effective of all the control strategies for the control and management of the burden of oncogenic HPV and TB co-infection.

## Data Availability

Not applicable

https://www.indexmundi.com/china/demographics\_profile

